# A data-driven regional amyloid PET score predicts cognitive decline beyond Centiloid

**DOI:** 10.64898/2026.08.26.26360248

**Authors:** Takumi Hirose, Wado Akamatsu, Tadafumi Kato, the Alzheimer’s Disease Neuroimaging Initiative

**Affiliations:** Center for Genomic and Regenerative Medicine, Juntendo University, Tokyo, Japan; Department of Psychiatry, Juntendo University, Tokyo, Japan

**Keywords:** Alzheimer’s disease, Positron-Emission Tomography, Amyloid beta-Peptides, Cognitive Dysfunction, Prognosis, Biomarkers

## Abstract

**Background:** The Centiloid (CL) scale standardizes global amyloid PET quantification and is widely used to define amyloid positivity. As a global summary measure, however, CL may not fully reflect the regional distribution of amyloid deposition, which can carry additional prognostic information about the rate of cognitive decline.

**Objective:** To develop and externally validate a fixed, regional amyloid PET composite score that complements CL for predicting cognitive decline in Alzheimer’s disease.

**Methods:** The Regional Amyloid PET Score (RAPS) was derived from 82 FreeSurfer regions using machine learning with bootstrap stability selection to predict the rate of change in CDR–Sum of Boxes (CDR-SB) in 433 amyloid-positive ADNI participants imaged with [^18^F]florbetapir (*n* = 357) or [^18^F]florbetaben (*n* = 76). The fixed nine-region weights were applied without retraining in a small ADNI [^18^F]florbetaben subgroup (*N* = 71, 68 of whom were in the discovery cohort; sensitivity analysis) and two external validation cohorts, NACC SCAN (*N* = 1531; four tracers) and OASIS-3 (*N* = 428).

**Results:** RAPS comprised nine regions. In ADNI post-selection evaluation, RAPS correlated more strongly with CDR-SB slope than CL and showed higher discrimination of rapid decliners (AUC 0.813 vs 0.708). RAPS remained associated with progression after CL adjustment in both external cohorts, although neither external AUC difference was significant. Cross-cohort meta-analysis including ADNI development and the two external cohorts favored RAPS alone over CL alone (pooled ΔAUC +0.069; *I*^2^ = 1.6%).

**Conclusions:** RAPS, a fixed regional amyloid PET–derived score, may complement CL for prognostic stratification in Alzheimer’s disease research.

## Introduction

Amyloid positron emission tomography (PET) has transformed the diagnosis and management of Alzheimer’s disease (AD) by enabling in vivo quantification of cerebral amyloid-β (Aβ) plaque burden.^1^ With the clinical introduction of anti-Aβ immunotherapies,^2–4^ confirmation of amyloid pathology has become essential for treatment eligibility; because these therapies carry a risk of amyloid-related imaging abnormalities (ARIA), accurate identification of individuals at greatest risk of near-term clinical progression is a pressing priority. Amyloid positivity defines Alzheimer’s pathologic change,^5^ yet not all amyloid-positive individuals show near-term cognitive progression, particularly at lower amyloid burdens.

The Centiloid (CL) scale standardized amyloid PET quantification across tracers and centers^6–8^ and is widely used in clinical trials and practice as a measure of global amyloid burden; amyloid PET itself is used clinically under appropriate-use criteria.^9^ However, the CL approach collapses the spatial distribution of amyloid into a single global metric, overlooking where amyloid is deposited—which may matter when global burden alone provides limited prognostic separation, including in the intermediate range.

Neuropathological studies have established distinct spatial progressions of neurofibrillary and amyloid pathology; Braak and Braak^10^ described neurofibrillary staging, whereas Thal et al.^11^ described five phases of Aβ spread from neocortical to allocortical, subcortical, and brainstem regions, and in vivo PET staging has confirmed analogous amyloid patterns.^12–14^ Tau PET has demonstrated strong prognostic value; Ossenkoppele et al.^15^ showed that baseline tau PET predicted subsequent cognitive decline and outperformed amyloid PET and structural MRI, raising the question of whether a region-specific approach could improve amyloid PET-based prognosis.

Despite growing evidence for regional amyloid quantification, existing studies have largely relied on global summary measures (CL or composite SUVRs) or individual a priori–selected ROIs. Few studies have systematically developed a sparse, interpretable regional amyloid PET composite optimized for longitudinal cognitive decline and externally evaluated across cohorts and tracers without retraining. Previous studies^16–18^ have examined regional amyloid accumulation, staging, and spatial distribution, using approaches that differ from a fixed regional composite optimized for cognitive decline.

In this study, we developed the Regional Amyloid PET Score (RAPS), a data-driven regional amyloid composite built from the Alzheimer’s Disease Neuroimaging Initiative (ADNI) cohort and externally evaluated in the National Alzheimer’s Coordinating Center (NACC) Standardized Centralized Alzheimer’s & Related Dementias Neuroimaging (SCAN) multi-tracer cohort and the Open Access Series of Imaging Studies (OASIS-3), with a sensitivity analysis in a small ADNI [^18^F]florbetaben subgroup that largely overlaps the discovery cohort. We tested whether this data-driven regional composite provides prognostic information for longitudinal cognitive decline complementary to global Centiloid—across amyloid burden strata, *APOE* ε4 genotype, and survival outcomes—with secondary exploratory analyses in the CL 20–60 subgroup.

## Methods

### Study Participants

#### Discovery cohort (ADNI)

Data used in the preparation of this article were obtained from the ADNI database (adni.loni.usc.edu).^19–20^ The ADNI was launched in 2003 as a public-private partnership, led by Principal Investigator Michael W. Weiner, MD. The primary goal of ADNI has been to test whether serial magnetic resonance imaging (MRI), positron emission tomography (PET), other biological markers, and clinical and neuropsychological assessment can be combined to measure the progression of mild cognitive impairment (MCI) and early Alzheimer’s disease (AD). Inclusion criteria were cognitively normal (CN) or MCI at ADNI enrollment, available amyloid PET with an [^18^F]-labeled tracer ([^18^F]florbetapir or [^18^F]florbetaben), *APOE* genotyping, baseline (earliest PET) Centiloid ≥ 20, and ≥ 2 years of longitudinal Clinical Dementia Rating (CDR) follow-up. From 5062 participants, 433 amyloid-positive CN/MCI individuals were included. The baseline amyloid PET scan (the earliest scan with a Centiloid value) was [^18^F]florbetapir in 357 participants and [^18^F]florbetaben in 76; both tracers were processed through the same UC Berkeley pipeline (below), with global burden expressed on the Centiloid scale, whereas the regional model inputs were unharmonized SUVRs. Although the Centiloid scale harmonizes global amyloid burden across tracers, regional amyloid PET uptake patterns can differ between tracers; regional SUVRs from the two tracers were pooled, and tracer was not included as a covariate or stratification factor at any stage of derivation (residualization, feature selection, standardization, or refitting). Baseline Centiloid was similar in the two tracer groups (74.2 ± 40.4 vs 73.4 ± 34.9). Performance across tracers was examined in the multi-tracer NACC SCAN validation cohort. The CL ≥ 20 restriction targeted prognostic stratification after amyloid positivity was established; it was not intended to develop a score for distinguishing amyloid-negative from amyloid-positive individuals.

#### Overlapping florbetaben subgroup (ADNI; sensitivity analysis)

A subgroup of ADNI participants (*N* = 71) with [^18^F]florbetaben (FBB) PET was used as a small-sample sensitivity analysis, applying the fixed RAPS weights without retraining. This subgroup was drawn from the same ADNI longitudinal dataset as the discovery cohort (participant-level CDR-SB slopes as in the discovery analysis) with Centiloid ≥ 20 at the FBB scan (source file *N* = 72); one participant with only a same-day CDR assessment and no subsequent assessment had zero follow-up time and was excluded (*N* = 71). The subgroup comprised largely the same individuals as the discovery cohort (68 of 71 overlapping, 67 of whom entered the discovery cohort through a florbetaben scan); it therefore provides only a descriptive check of whether the fixed weights remain informative in a small florbetaben-defined subgroup and, because 68 of 71 participants contributed to model development, provides no independent evidence of predictive performance or tracer transportability. For each participant, the imaging index was the earliest FBB scan with all nine RAPS regions available, and Centiloid was taken from that scan; for the Cox analyses, time zero was the date of that scan, whereas the CDR-SB slope used for the correlation and ROC analyses was the participant-level slope from the longitudinal CDR-SB series and was not recomputed relative to the FBB index date. The correlation and ROC analyses should therefore not be interpreted as prediction of decline exclusively after that scan.

#### External validation cohorts

Two external validation cohorts were used. The NACC SCAN initiative^21–22^ provided 1707 participants imaged with four amyloid PET tracers ([^18^F]florbetapir, [^18^F]florbetaben, [^11^C]PiB, [^18^F]NAV4694); inclusion criteria and processing are described below (External Validation) and in Supplementary Methods. OASIS-3 (Washington University, St. Louis)^23^ provided 429 participants with [^18^F]florbetapir PET and longitudinal CDR data; after excluding one with missing Centiloid, the analytic cohort comprised 428; CL strata (CL ≥ 20; CL 20–60 and CL ≥ 60) were used in sensitivity analyses.

#### Amyloid PET Acquisition and Processing

In ADNI, regional standardized uptake value ratios (SUVRs) were calculated by the UC Berkeley Jagust Laboratory^24–25^ using FreeSurfer (version 7.1)^26^ parcellation (Desikan-Killiany cortical regions and subcortical regions from automated segmentation [aseg]),^27^ with whole cerebellum as the reference region for normalization;^24^ Centiloid was derived using tracer- and reference-region-specific conversions.^28^ All 82 ROIs were retained (68 cortical, 14 subcortical). No partial volume effect (PVE) correction was applied.^29^ Visualizations used MRIcroGL.^30^ Amyloid positivity was defined as a global CL ≥ 20. This threshold approximates the neuropathological level at which amyloid PET discriminates moderate-to-frequent neuritic plaques (≈19 CL for florbetaben^31^) and equals the ADNI florbetapir positivity threshold of whole-cerebellum-normalized SUVR 1.11 (= 20 CL^28^); comparable positivity cutoffs (≈19 CL) have been adopted in independent cohorts,^32^ and CL ≤ 20 corresponds to the amyloid-negative boundary in a recent quantification study.^33^

#### Cognitive Outcomes

In ADNI, the primary cognitive outcome was the individual rate of change in the Clinical Dementia Rating Sum of Boxes (CDR-SB),^34–35^ estimated by linear mixed-effects models (random intercepts and slopes per participant; details in Supplementary Methods). The decline rate was the per-participant slope (best linear unbiased predictor, BLUP), with ordinary least squares (OLS) slopes used in a sensitivity comparison. The original repeat-observation check counted post-origin data rows, including rows with missing CDR-SB, instead of counting nonmissing CDR-SB assessments as intended. Thirteen of the 433 participants had one retained CDR-SB observation; their BLUP slopes were estimated using the population model rather than directly observed within-person change. In the corrected evaluation, CDR dates were taken from the clinical assessment records and identical repeated observations were counted once; the fixed RAPS model and saved out-of-fold scores were retained. Model-development results (region selection, coefficients and internal cross-validation) are retained from the original development analysis; subsequent evaluations of the fixed scores use the corrected clinical data. The participant-level ADNI slopes were estimated from the retained longitudinal clinical series and do not necessarily represent decline exclusively after the index PET scan. External-cohort slope methods and observation windows are described in Supplementary Methods.

#### RAPS Construction and Post-selection Cross-Validation

The Regional Amyloid PET Score (RAPS) was constructed in the ADNI discovery cohort from 82 FreeSurfer regions (68 Desikan-Killiany cortical regions and 14 aseg-derived subcortical regions; Supplementary Table S1) using an outer-fold cross-validation framework^36^ to restrict model fitting and hyperparameter selection to the training data. Within each outer fold, covariate regression (age, sex, education, *APOE* ε4 count, baseline CDR-SB; Centiloid excluded to enable RAPS vs CL comparison) was fitted on the training set to residualize the CDR-SB slope and applied to the held-out test set. Training-set–standardized regional SUVRs were entered into an elastic net regression^37–38^ (ElasticNetCV, 5-fold inner CV; L1 ratio ∈ {0.1–0.9}; α auto-optimized). Out-of-fold predictions were concatenated and Z-standardized to yield post-selection out-of-fold RAPS scores, and a final fixed-coefficient model was refitted on the complete dataset for external validation. The definitive RAPS was the intersection of non-zero ElasticNet coefficients and bootstrap selection frequency ≥ 80%, serialized without retraining (Supplementary Methods). Because the nine regions were selected on the full discovery sample, the 9-region out-of-fold metrics reflect a post-selection cross-validation in which coefficients and hyperparameters—but not the choice of regions—were re-estimated within each fold, and may be optimistic for the feature-selection step; the comparison with the fully nested 82-region model should be interpreted with this asymmetry in mind.

ROI selection stability was assessed by 1000 bootstrap iterations (resampling with replacement, refitting elastic net de novo); ROIs selected in ≥80% were bootstrap-stable. The final RAPS was the intersection of bootstrap-stable and non-zero-coefficient ROIs. OLS-derived and BLUP-derived CDR-SB slopes were highly correlated (Supplementary Figure S1).

As a sensitivity analysis, the original eligibility sequence was retained with baseline CL > 0, and the same 82-region, 5×5 cross-validation, 1000-bootstrap and 80% intersection procedure was repeated. The resulting model was compared directly with the locked nine-region RAPS in the same external participants without external retraining. Corrected scan-level inputs and slopes re-estimated in an expanded LME pool were used for sensitivity retraining. A CL ≥ 20 model was also retrained using the same processing and the same slopes for shared participants, to assess the development-range difference. The original nine-region RAPS and corrected primary evaluations were retained (Supplementary Methods). We also examined exclusion of the 13 participants with one retained CDR-SB observation by refitting the original nine-region set and by repeating region selection in the remaining 420 participants (Supplementary Methods).

#### Statistical Analysis

RAPS vs CL performance was evaluated with Steiger tests^39^ for dependent correlations and paired bootstrap tests^40^ for areas under the curve (AUCs) at CDR-SB slope thresholds > 0.5, > 1.0, and > 1.5/yr. Categorical net reclassification improvement (NRI; tertile cutoffs) and integrated discrimination improvement (IDI)^41^ used 5-fold cross-validated logistic-regression probabilities (Centiloid vs Centiloid + RAPS) at the same thresholds. Incremental prognostic value used nested Cox models with 5-fold CV concordance. NACC Cox used visit-level CDR-SB (event = first post-PET CDR-SB ≥ baseline + 1.0; 178 events/1531).

Subgroup analyses examined RAPS performance across CL stages (early: CL 20–60 vs late: CL ≥60), *APOE* ε4 carrier status, sex, and baseline cognitive status (CDR-SB = 0 vs > 0), with CL threshold sensitivity at 50, 60, and 70; these CL stage boundaries were pre-specified for exploratory subgroup analysis rather than as diagnostic thresholds, where CL 20 defines amyloid positivity and the CL 20–60 window was examined as a low-to-moderate amyloid-burden stratum. *APOE* × RAPS interaction was tested by OLS regression.

Time-to-event analysis used Kaplan–Meier and Cox proportional hazards regression.^42^ Progression was the first post-baseline visit with CDR-SB ≥ baseline + 1.0; otherwise participants were censored at last assessment. Cox models were an age- and sex-adjusted RAPS model (M1), M1 plus Centiloid (M2), and a fully adjusted joint model (M3: RAPS, CL, age, sex, education, *APOE* ε4 count, baseline CDR-SB); in the ADNI florbetaben subgroup, sex was omitted from all three models and education from M3 because of the small sample size. Discrimination over follow-up used inverse probability of censoring weighting (IPCW) cumulative/dynamic time-dependent AUCs at 1–8-year horizons.^43–44^ The proportional hazards assumption was tested with Schoenfeld residuals.

An exploratory analysis stratified the CL 20–60 subgroup into RAPS-high and RAPS-low groups by median split within the CL 20–60 subgroup, computed separately in ADNI and OASIS-3. Cognitive decline rates, rapid-decliner proportions, and ROI-level SUVR patterns were compared.

Cross-cohort synthesis. ΔAUC (RAPS alone − Centiloid alone) at slope > 1.0/year and M3 log hazard ratios were pooled by random-effects (DerSimonian-Laird) meta-analysis; heterogeneity was assessed by Cochran’s *Q* and *I*^2^. Primary pooling included the three independent cohorts (ADNI discovery, NACC SCAN, and OASIS-3); the overlapping ADNI FBB subgroup was excluded from pooling and evaluated separately as a descriptive sensitivity analysis.

The corrected evaluations used Python 3.12 and R 4.5.2 (package versions in Supplementary Methods); Supplementary Table S2 records the original model-development settings. Two-sided *P* < .05 was significant. Benjamini-Hochberg FDR correction was applied within the subgroup correlation family (9 subgroups); baseline-diagnosis (CN/MCI) subgroup analyses were exploratory and uncorrected. Pre-specified primary comparisons were Steiger’s test and the paired bootstrap AUC comparison at the > 1.0/year threshold; additional thresholds were supportive and uncorrected; NRI/IDI estimates were exploratory and descriptive, without significance testing.

#### External Validation

The fixed 9-ROI RAPS weights were applied to the NACC^21^ SCAN^22^ cohort. The source sample comprised 1707 participants from 37 NIA-funded Alzheimer’s Disease Research Centers, imaged with four tracers: [^18^F]florbetapir (*n* = 619), [^18^F]florbetaben (*n* = 280), [^11^C]PiB (*n* = 709), or [^18^F]NAV4694 (*n* = 99); demographics in Table 1. Regional SUVRs were used as supplied by SCAN, referenced to whole cerebellum. The primary analytic cohort was Centiloid-unrestricted (*N* = 1531; 178 visit-level Cox events; 117 rapid-decliner events at slope > 1.0/year; derivation in Results). Of these participants, 539 had Centiloid ≥ 20. Cox models, paired bootstrap AUC comparisons, and Pencina-style reclassification metrics were used. The CL ≥ 20 subset was also evaluated using the existing Cox covariates, clinical definitions and full-cohort predictor standardization (Supplementary Methods).

**Table 1.**
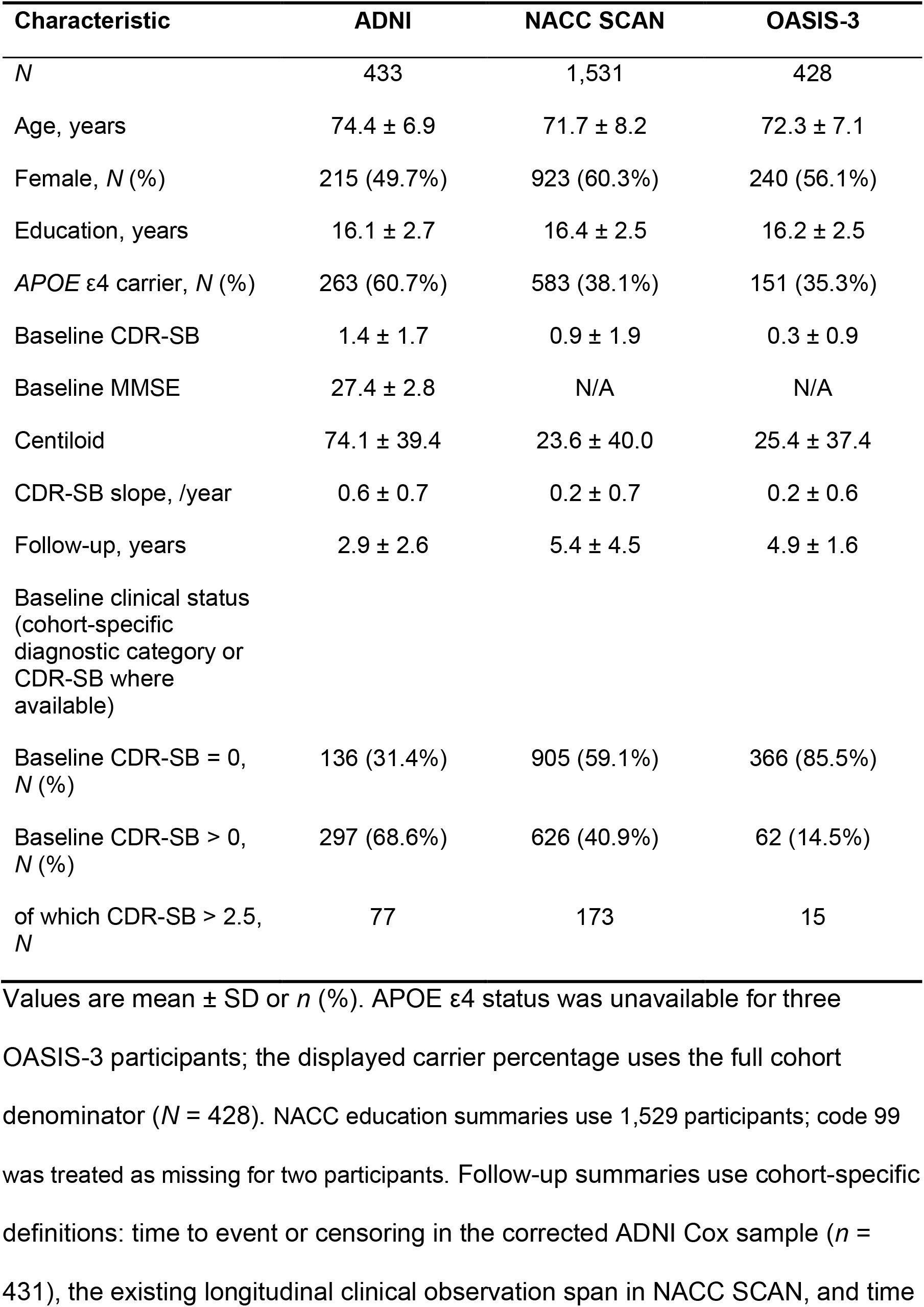
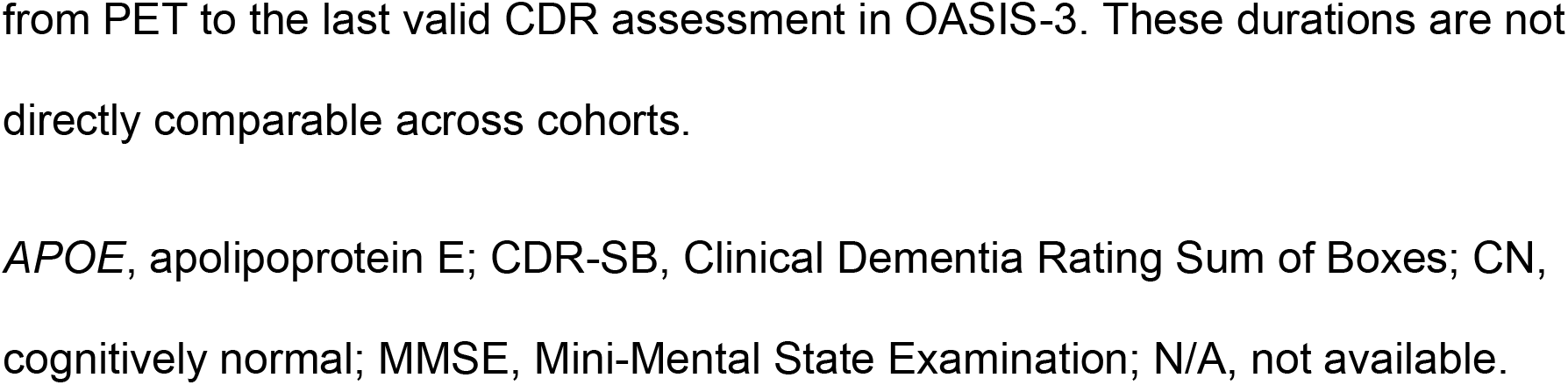
Demographic and Clinical Characteristics of Study Cohorts.

| Characteristic | ADNI | NACC SCAN | OASIS-3 |
|---|---|---|---|
| *N* | 433 | 1,531 | 428 |
| Age, years | 74.4 ± 6.9 | 71.7 ± 8.2 | 72.3 ± 7.1 |
| Female, *N* (%) | 215 (49.7%) | 923 (60.3%) | 240 (56.1%) |
| Education, years | 16.1 ± 2.7 | 16.4 ± 2.5 | 16.2 ± 2.5 |
| *APOE* ε4 carrier, *N* (%) | 263 (60.7%) | 583 (38.1%) | 151 (35.3%) |
| Baseline CDR-SB | 1.4 ± 1.7 | 0.9 ± 1.9 | 0.3 ± 0.9 |
| Baseline MMSE | 27.4 ± 2.8 | N/A | N/A |
| Centiloid | 74.1 ± 39.4 | 23.6 ± 40.0 | 25.4 ± 37.4 |
| CDR-SB slope, /year | 0.6 ± 0.7 | 0.2 ± 0.7 | 0.2 ± 0.6 |
| Follow-up, years | 2.9 ± 2.6 | 5.4 ± 4.5 | 4.9 ± 1.6 |
| Baseline clinical status (cohort-specific diagnostic category or CDR-SB where available) |  |  |  |
| Baseline CDR-SB = 0, *N* (%) | 136 (31.4%) | 905 (59.1%) | 366 (85.5%) |
| Baseline CDR-SB > 0, *N* (%) | 297 (68.6%) | 626 (40.9%) | 62 (14.5%) |
| of which CDR-SB > 2.5, *N* | 77 | 173 | 15 |

Values are mean ± SD or *n* (%). APOE ε4 status was unavailable for three OASIS-3 participants; the displayed carrier percentage uses the full cohort denominator (*N* = 428). NACC education summaries use 1,529 participants; code 99 was treated as missing for two participants. Follow-up summaries use cohort-specific definitions: time to event or censoring in the corrected ADNI Cox sample (*n* = 431), the existing longitudinal clinical observation span in NACC SCAN, and time from PET to the last valid CDR assessment in OASIS-3. These durations are not directly comparable across cohorts.
*APOE*, apolipoprotein E; CDR-SB, Clinical Dementia Rating Sum of Boxes; CN, cognitively normal; MMSE, Mini-Mental State Examination; N/A, not available.

RAPS was similarly applied to OASIS-3 without retraining (Supplementary Methods). For three participants without a matching entry in the Centiloid table, Centiloid was calculated from the same-scan non-PVC mean cortical SUVR using the OASIS-3 conversion equation (163.6 × SUVR − 181.0).

An exploratory ADNI tau-adjustment analysis required the original amyloid PET and the nearest available non-PVC meta-temporal flortaucipir PET measurement to be within one year. For this ancillary analysis, CDR-SB slopes were re-estimated, using at least two CDR observations after both scans and a final observation at least two years after the later scan. Linear models compared the association of fixed post-selection out-of-fold RAPS with these slopes before and after tau adjustment in identical complete cases, additionally adjusting for CL, age, sex, education, *APOE* ε4 count and the first CDR-SB after this ancillary index (Supplementary Methods).

## Results

### Cohort Characteristics

From 5062 ADNI participants, inclusion criteria yielded 433 amyloid-positive CN or MCI individuals for the discovery cohort (Figure 1). Demographic and clinical characteristics are summarized in Table 1 (mean baseline CL 74.1; 60.7% *APOE* ε4 carriers). The mean LME-estimated CDR-SB slope was 0.58 points per year (SD 0.73), and 96 participants (22.2%) were rapid decliners (slope > 1.0).

**Figure 1.**
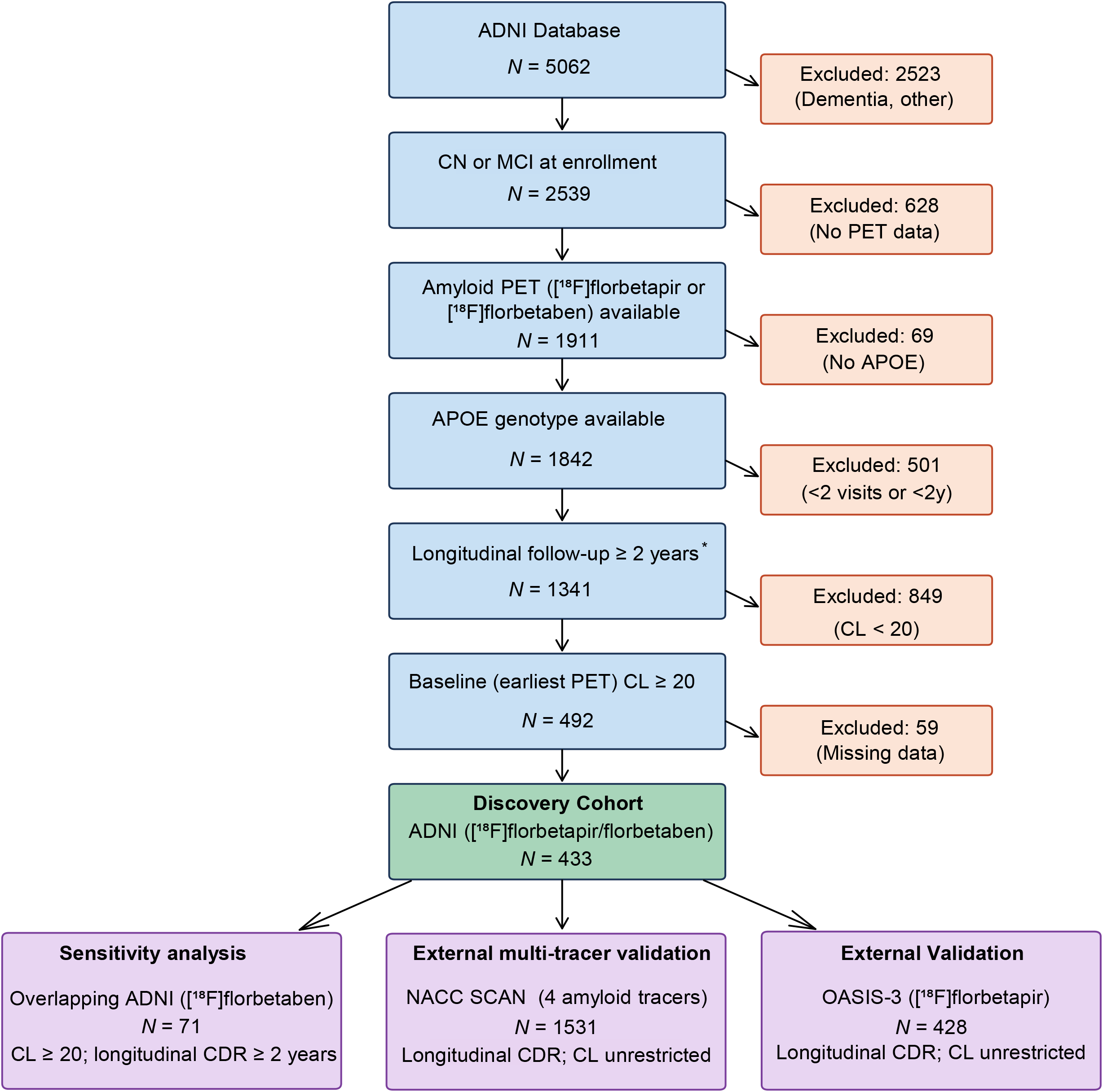
Study participant flow diagram. Sequential application of inclusion and exclusion criteria yielded the ADNI discovery cohort ([^18^F]florbetapir *n* = 357, [^18^F]florbetaben *n* = 76; *N* = 433 from 5062), the overlapping ADNI [^18^F]florbetaben subgroup used for sensitivity analysis (*N* = 71), the NACC SCAN multi-tracer source cohort (four tracers; *N* = 1707; primary analytic *N* = 1531), and the OASIS-3 external validation cohort ([^18^F]florbetapir; *N* = 428). Longitudinal CDR follow-up was required across cohorts. *The ADNI follow-up criterion was applied to the original clinical records; 13 of the 433 participants had only one CDR-SB observation retained for slope estimation and received population-model-based BLUP slopes. In ADNI, amyloid positivity was defined as baseline (earliest PET) Centiloid ≥ 20; CL ≥ 20 also defined the NACC SCAN amyloid-positive subset. NACC SCAN and OASIS-3 were used as CL-unrestricted external validation cohorts, with CL strata examined only in sensitivity analyses.

The OASIS-3 external validation cohort comprised 429 participants with [^18^F]florbetapir PET and longitudinal CDR data; one was excluded for missing Centiloid, yielding *N* = 428. Demographic and clinical characteristics of the ADNI discovery, NACC SCAN, and OASIS-3 cohorts are presented in Table 1.

### RAPS Construction and Internal Validation

In the original development analysis, post-selection internal validation used 5×5 cross-validation predicting covariate-adjusted CDR-SB slope (mean fold *R*^2^ = 0.090, mean fold Pearson *r* = 0.381; Supplementary Figure S2). Applied to all 82 ROIs (*N* = 433), ElasticNet selected 15 non-zero ROIs; 9 also met bootstrap stability ≥ 80%, constituting the final RAPS. In the 9-ROI refit, the left hippocampus had the largest absolute coefficient (β = −0.169); the remaining eight cortical and subcortical regions are shown in Figure 2A and Supplementary Table S3. OLS-derived and BLUP-derived slopes showed a strong correlation (*r* = 0.919; Supplementary Figure S1).

**Figure 2.**
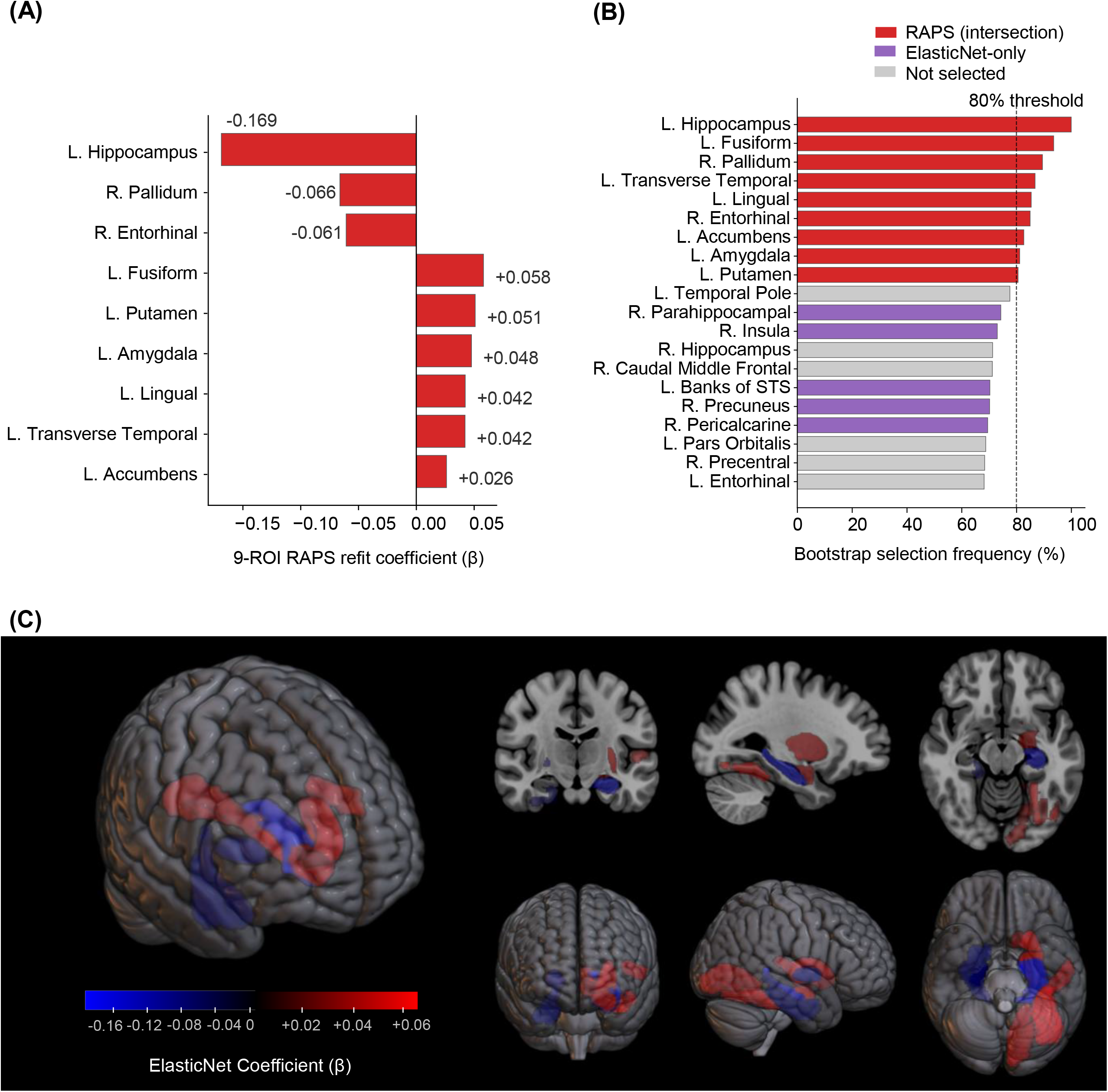
RAPS ROI Selection and Spatial Distribution. (A) Final 9-ROI RAPS refit coefficients (α = 0.058, L1 ratio = 0.10), sorted by absolute magnitude. Numerical β values are annotated next to each bar; positive values indicate that higher SUVR is associated with faster cognitive decline. (B) Bootstrap selection frequency (1000 iterations) for the top 20 ROIs. The 9 RAPS ROIs (red; intersection of bootstrap ≥ 80% and ElasticNet non-zero); ElasticNet-only ROIs (purple); remaining ROIs (gray). The 80% stability threshold is indicated by a dashed line. (C) Three-dimensional brain rendering (MRIcroGL) of the 9 RAPS ROIs with axial, sagittal, and coronal views. Red indicates positive coefficients (cortical and subcortical regions predicting faster decline); blue indicates negative coefficients (hippocampus, pallidum, entorhinal cortex). Original model-development results, retained without refitting after the data audit.

In the original post-selection analysis, fold-level *R*² ranged from −0.08 to 0.21, and the pooled out-of-fold correlation was 0.363 (*P* < .001; Supplementary Figure S2).

### Bootstrap Stability and 9-ROI RAPS Definition

Bootstrap resampling (1000 iterations) identified the 9 ROIs meeting both criteria (selection ≥ 80% and non-zero ElasticNet coefficient; Figure 2B; Supplementary Figure S3). The left hippocampus was selected in 100% of iterations with perfect sign consistency. Several stable ROIs have 95% confidence intervals crossing zero (Supplementary Figure S3), indicating that RAPS’s predictive value arises from the combined 9-ROI pattern rather than any single region.

The 9-ROI RAPS score—the intersection of ElasticNet-selected and bootstrap-stable ROIs—was retained as the final model (Figure 2C). In the post-selection comparison, the fixed 9-ROI model showed slightly higher performance than the fully nested 82-ROI model (mean fold *r* = 0.381 vs 0.351; Supplementary Figure S4).

### RAPS vs Centiloid

RAPS showed stronger zero-order correlation with CDR-SB slope than global CL (*r* = 0.545 vs 0.314; Steiger *z* = 5.27, *P* < .001; Figure 3A,B). The RAPS–CL correlation was moderate (*r* = 0.445; Supplementary Table S4). In discriminating rapid cognitive decliners, RAPS outperformed CL across all CDR-SB slope thresholds, with a significant gain in discrimination at the primary > 1.0/year threshold (bootstrap *P* = .001; Table 2, Panel A; Figure 3C–E). NRI and IDI were positive in exploratory descriptive analyses (Table 2, Panel B). RAPS also exceeded left hippocampus SUVR alone (AUC = 0.666), supporting multi-ROI weighting.

**Figure 3.**
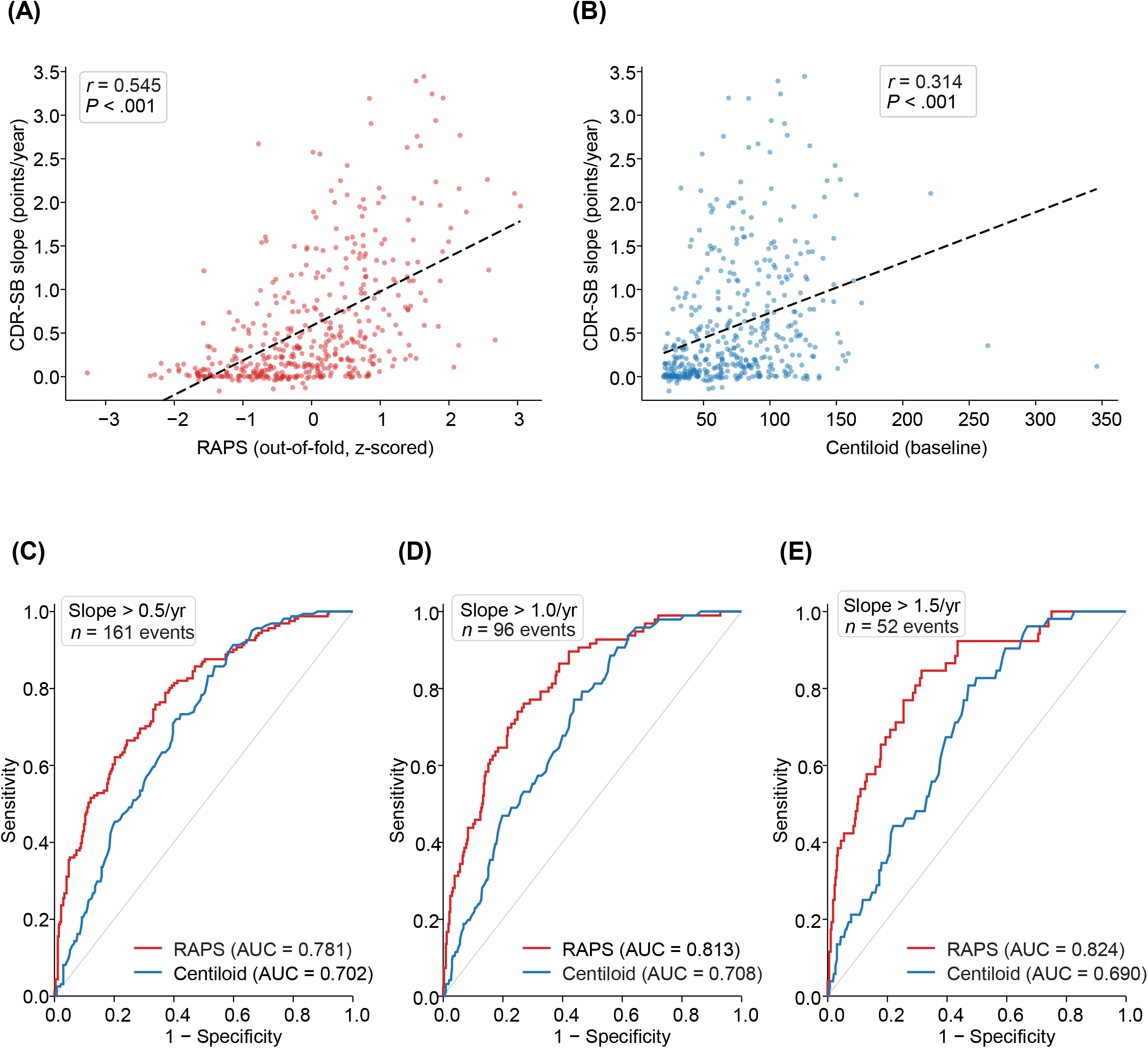
RAPS Outperforms Centiloid in ADNI Post-selection Evaluation of CDR-SB Progression (*N* = 433). (A) Scatter plot of RAPS (out-of-fold, z-scored) versus CDR-SB slope (Pearson *r* = 0.545, *P* < .001). (B) Scatter plot of baseline Centiloid versus CDR-SB slope (Pearson *r* = 0.314, *P* < .001). Steiger test for dependent correlations confirms RAPS is significantly stronger than CL (*z* = 5.27, *P* < .001). Dashed lines indicate linear regression fits. (C–E) Receiver operating characteristic curves comparing RAPS and Centiloid for predicting rapid cognitive decline at three thresholds: CDR-SB slope > 0.5 (C), > 1.0 (D), and > 1.5 (E) points per year. AUC values shown for each curve.

**Table 2.** RAPS vs Centiloid: Post-selection Out-of-Fold Predictive Performance in the ADNI Discovery Cohort.

| **Panel A: ROC Analysis** |  |  |  |  |  |
|---|---|---|---|---|---|
| **Threshold (CDR-SB/yr)** | ***N* rapid** | **RAPS AUC** | **CL AUC** | **ΔAUC** | **Bootstrap *P*** |
| > 0.5 | 161 | 0.781 | 0.702 | +0.079 | .006 |
| > 1.0 | 96 | 0.813 | 0.708 | +0.105 | .001 |
| > 1.5 | 52 | 0.824 | 0.690 | +0.134 | .003 |
| ***N*** | ***r* (RAPS)** | ***r* (CL)** | **Δ*r*** | **Steiger *z*** | ***P*** |
| 433 | 0.545 | 0.314 | 0.232 | 5.27 | < .001 |
| **Panel B: Exploratory Reclassification Metrics** |  |  |  |  |  |
| **Threshold (CDR-SB/yr)** | **NRI** | **IDI** |  |  |  |
| > 0.5 | 0.236 | 0.147 |  |  |  |
| *> 1.0* | *0.325* | *0.169* |  |  |  |
| > 1.5 | 0.484 | 0.152 |  |  |  |
| **Panel C: Cox Proportional Hazards Models** |  |  |  |  |  |
| **Model / Covariate** | **HR** | **95% CI** | ***P*** |  |  |
| RAPS model (RAPS + age + sex) |  |  |  |  |  |
| RAPS (z) | 2.12 | (1.81–2.47) | < .001 |  |  |
| CL model (CL + age + sex) |  |  |  |  |  |
| CL (z) | 1.45 | (1.30–1.62) | < .001 |  |  |
| Joint multivariable model (RAPS + CL + age + sex + education + APOE ε4 + baseline CDR-SB) |  |  |  |  |  |
| RAPS (z) | 1.61 | (1.35–1.92) | < .001 |  |  |
| CL (z) | 1.28 | (1.10–1.50) | .002 |  |  |
| Age (per year) | 0.98 | (0.96–1.01) | .155 |  |  |
| Female sex | 1.01 | (0.75–1.35) | .970 |  |  |
| Education (per year) | 1.01 | (0.95–1.06) | .861 |  |  |
| APOE ε4 (per allele) | 1.07 | (0.85–1.34) | .578 |  |  |
| Baseline CDR-SB (per point) | 1.28 | (1.19–1.39) | < .001 |  |  |

Out-of-fold predictions from 5×5 post-selection cross-validation in the ADNI discovery cohort (*N* = 433). Rapid decline is defined by the indicated annual CDR-SB progression threshold; *N* rapid denotes the number of progressors. AUCs are compared with a paired bootstrap test and the Steiger test compares dependent correlations. Panel C presents multivariable Cox proportional hazards models (*N* = 431, 190 events). The nine-region feature set was selected using the full discovery cohort; coefficients and hyperparameters were re-estimated within each outer fold. Panel B reports exploratory, descriptive NRI and IDI estimates; *P* values are not reported.
APOE, apolipoprotein E; AUC, area under the receiver operating characteristic curve; CDR-SB, Clinical Dementia Rating Sum of Boxes; CI, confidence interval; CL, Centiloid; HR, hazard ratio; IDI, integrated discrimination improvement; NRI, net reclassification improvement; RAPS, Regional Amyloid PET Score; ΔAUC, difference in AUC (RAPS − CL); Δ*r*, difference in correlation.

### Subgroup Analyses

RAPS correlated with CDR-SB slope across all subgroups examined (Supplementary Figure S5A; Supplementary Table S5). However, performance was weaker in two clinically important subgroups: participants with baseline CDR-SB = 0 (*r* = 0.246, *n* = 136) and the high-amyloid stratum (CL ≥ 60; *r* = 0.473, *n* = 260). These zero-order values are not directly comparable to the covariate-adjusted out-of-fold *r* (0.363) because their outcomes were not residualized for covariates and were evaluated using the corrected, rather than original-development, clinical outcome data. The CN/MCI diagnostic strata were defined by diagnosis at ADNI enrollment.

A significant *APOE* ε4 × RAPS interaction was observed (β-interaction = 0.074, *P* = .047; Supplementary Table S6). CL-stratified analysis revealed distinct ROI weight patterns: the left hippocampus dominated in the late stage (β = −0.213), whereas in the early stage predictive information was distributed across broader cortical regions (Supplementary Figure S5B). Sensitivity analyses across CL thresholds (50, 60, 70) showed stable RAPS performance in both strata (early-stage *r* = 0.494–0.551). The *APOE* ε4 interaction was exploratory and was not adjusted across all interaction tests.

Because cognitively normal and MCI participants may differ in their capacity for measurable decline, we analyzed the two diagnostic groups defined at ADNI enrollment separately. RAPS predicted CDR-SB slope in both CN (*n* = 169, *r* = 0.427 [95% CI 0.295–0.543]) and MCI (*n* = 264, *r* = 0.526 [0.433–0.608]) participants and significantly outperformed Centiloid in each stratum (Steiger *P* = .003 and *P* < .001, respectively). RAPS remained an independent predictor of clinical progression after Centiloid adjustment in both groups (CN: M3 hazard ratio per SD = 2.60 [1.51–4.47]; MCI: 1.36 [1.13–1.65]). Rapid decliners (CDR-SB slope > 1.0/year) were infrequent among CN participants (8 events), so the discrimination AUC in this subgroup is exploratory; RAPS nonetheless retained prognostic value in time-to-event analysis (42 events).

### Time-to-event Prognostic Analysis

IPCW time-dependent AUC was numerically higher for RAPS than CL at early-to-mid horizons (mean 1–8 years: 0.774 vs 0.722; Supplementary Figure S5C), with the largest gap at 1–4 years. Median-split survival analysis showed survival separation for both RAPS and CL in the same analytic sample (*N* = 431) (RAPS hazard ratio [HR] = 3.86; CL HR = 2.58; both *P* < .001; Supplementary Figure S6), with median time to progression 2.1 years in RAPS-high versus 8.2 years in RAPS-low.

In Cox proportional hazards analysis (190 events in 431 participants), RAPS strongly predicted decline (HR = 2.12 per SD, 95% CI 1.81–2.47; concordance 0.705), outperforming CL alone (concordance 0.638). In the multivariable model with both scores plus demographic covariates, RAPS remained an independent predictor (HR = 1.61, 95% CI 1.35–1.92; Supplementary Figure S5D), as did baseline CDR-SB and CL (Table 2, Panel C; concordance 0.745; Supplementary Table S7).^45^

Schoenfeld testing supported the proportional hazards assumption for RAPS (*P* = .70); no violation was detected for Centiloid (*P* = .16), whereas baseline CDR-SB showed evidence of nonproportional hazards (*P* = .03).

RAPS yielded larger incremental concordance than CL in nested Cox models (5-fold CV C-index: clinical 0.700, +CL 0.711, +RAPS 0.740, +CL+RAPS 0.734); adding CL to clinical + RAPS did not improve CV concordance.

In an exploratory ADNI CL 20–60 subgroup (*n* = 173; Supplementary Figure S7), RAPS-based median stratification yielded a large effect on cognitive decline (RAPS-high vs RAPS-low CDR-SB slope 0.528 vs 0.092; Cohen’s *d* = 0.947), with higher discriminative AUC than Centiloid (0.877 vs 0.805 at > 1.0/yr). As a continuous variable, RAPS retained a strong partial correlation with slope after CL adjustment (partial *r* = 0.460, *P* < .001), and quartile stratification revealed a dose-response gradient (rapid decliners 0% Q1 → 30.2% Q4).

### Sensitivity Analysis in the Overlapping ADNI Florbetaben Subgroup

As a small-sample sensitivity analysis, the fixed 9-ROI RAPS weights were applied without retraining to the ADNI florbetaben subgroup (*N* = 71; mean Centiloid 69.9), which largely overlaps the discovery cohort. RAPS showed numerically higher discrimination of rapid decliners (AUC 0.885 vs 0.781; bootstrap *P* = .325; Figure 4A; Supplementary Table S8) and a stronger correlation with CDR-SB slope than CL (*r* = 0.556 vs 0.390; Steiger *P* = .095). Because of the overlap with the discovery cohort, these estimates are not independent and are reported for completeness.

**Figure 4.**
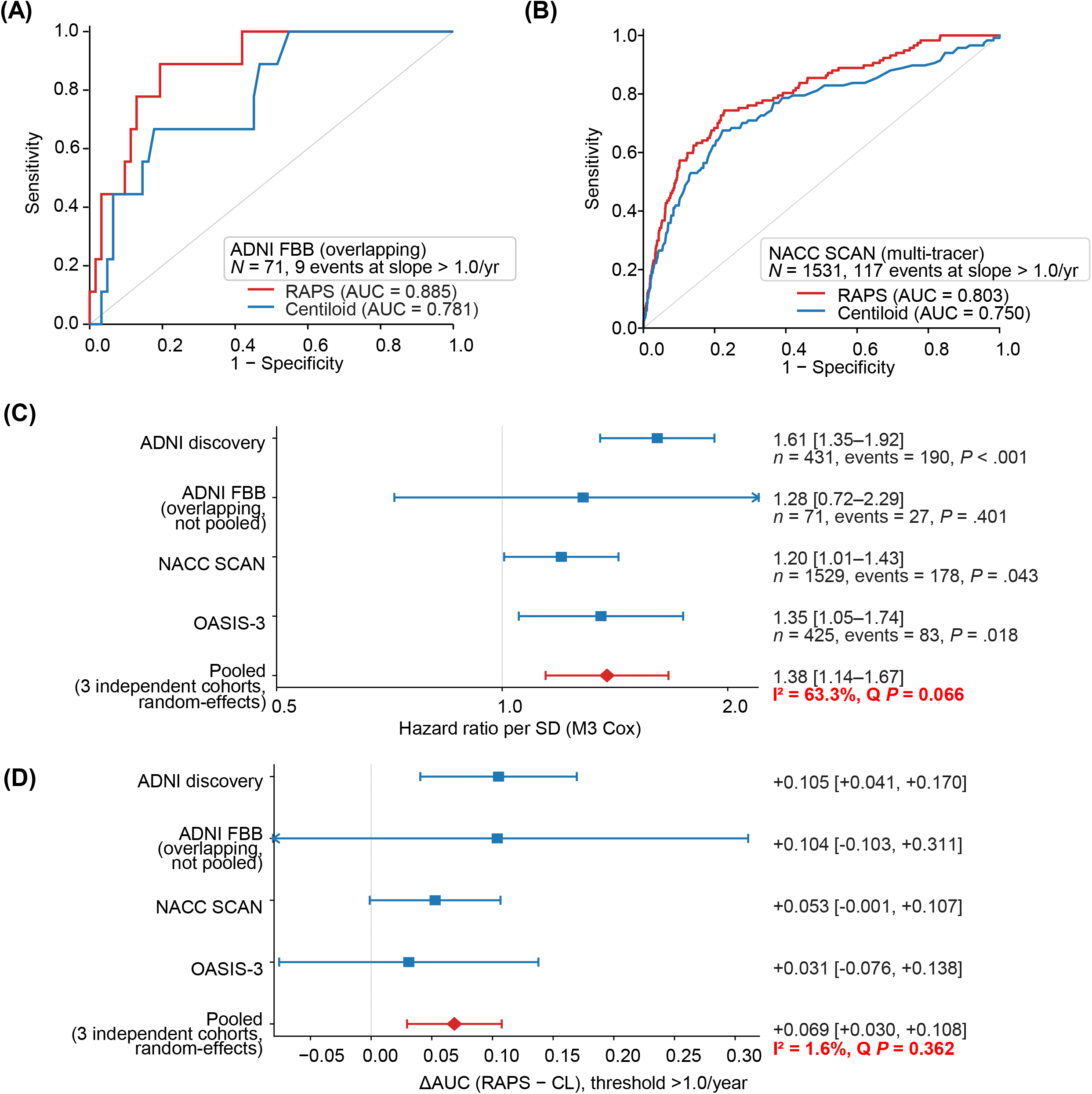
Multi-Cohort Evaluation and Cross-Cohort Synthesis of RAPS. (A) ROC curves for predicting rapid cognitive decline (CDR-SB slope > 1.0/year) in the overlapping ADNI [^18^F]florbetaben subgroup (*N* = 71, events = 9 at slope > 1.0/yr; sensitivity analysis). (B) ROC curves in the NACC SCAN analytic cohort (*N* = 1531, events = 117 at slope > 1.0/yr), showing overall RAPS performance across four amyloid tracers ([^18^F]florbetapir, [^18^F]florbetaben, [^11^C]PiB, [^18^F]NAV4694). (C) Forest plot of RAPS hazard ratios per SD from cohort-specific multivariable (M3) Cox models (adjusted for Centiloid and available clinical covariates per cohort; see Supplementary Table S7 for full specifications and Supplementary Table S10 for ΔAUC meta-analysis details). Hazard ratios were directionally consistent across cohort-specific M3 models (range 1.20–1.61); the primary ADNI Cox (with CDR-SB; HR = 1.61) is shown in Supplementary Figure S5D. The ADNI [^18^F]florbetaben models omitted sex (M1–M3) and education (M3) because of the small sample size (*N* = 71, 27 events); ADNI discovery values are out-of-fold RAPS predictions from post-selection 5×5 cross-validation. The right-pointing arrow indicates that the upper 95% confidence limit for the FBB subgroup extends beyond the plotted axis range. (D) Forest plot of ΔAUC (RAPS alone − Centiloid alone) for predicting CDR-SB slope > 1.0/year. Pooled estimate (filled red diamond) computed by random-effects (DerSimonian-Laird) meta-analysis across the three independent cohorts; heterogeneity *I*^2^ = 1.6%, Cochran *Q* = 2.03 (*df* = 2), *P* = .362. Event counts in panels A and B refer to slope-defined rapid decline, whereas those in panel C refer to visit-level increases in CDR-SB. Pooled estimates are from random-effects meta-analysis of the three independent cohorts (ADNI, NACC SCAN, OASIS-3); the ADNI FBB subgroup, which largely overlaps the discovery cohort, is displayed as a sensitivity analysis but excluded from the primary pooling. The left-pointing arrow in panel D indicates that the lower 95% confidence limit for the overlapping FBB subgroup extends beyond the plotted axis range.

### Multi-Tracer External Validation in NACC SCAN

The NACC SCAN source cohort comprised 1707 participants; excluding 167 without post-PET CDR follow-up and 9 same-day-only visits yielded the 1531-participant analytic cohort. RAPS correlated strongly with global Centiloid (*r* = 0.59). For rapid decline (slope > 1.0/yr; *N* = 1531; 117 events), RAPS AUC = 0.803 vs CL 0.750 (ΔAUC = +0.053; bootstrap *P* = .055; Figure 4B).

In the NACC SCAN analytic cohort (*N* = 1531; 178 visit-level events), RAPS and CL were each independent multivariable Cox predictors (M3, *N* = 1529 after excluding two missing education values: RAPS HR/SD = 1.20 [1.01–1.43], *P* = .043; CL HR = 1.31, *P* = .001; Figure 4C). Cross-tracer M1 HRs per SD were directionally consistent across four tracers (Supplementary Figure S8; tracer-stratified correlations with slope in Supplementary Table S9). RAPS improved reclassification at the primary threshold (categorical NRI = 0.230, IDI = 0.045; Supplementary Figure S9) and remained associated with progression in the CL 20–60 stratum (M1 HR = 1.49 [1.02–2.19], *P* = .041; 25 events/241).

### Cross-Cohort Synthesis

Across cohorts, M3 Cox RAPS HRs were directionally consistent (range 1.20–1.61 per SD; Figure 4C). Among the three independent cohorts, heterogeneity was substantial (pooled HR = 1.38, 95% CI 1.14–1.67; *I*^2^ = 63.3%), driven by the higher ADNI development-cohort estimate, based on post-selection out-of-fold RAPS scores and potentially retaining feature-selection optimism, whereas the two external cohort estimates were consistent (1.20–1.35). The difference in discrimination between RAPS alone and CL alone (ΔAUC at > 1.0/year) was directionally consistent across all four cohort datasets, significant in ADNI (+0.105, *P* = .001), not statistically significant in NACC (+0.053, *P* = .055), and nonsignificant in OASIS-3 (+0.031) and the overlapping ADNI FBB subgroup (+0.104; Figure 4D). Random-effects meta-analysis of the three independent cohorts yielded pooled ΔAUC = +0.069 (95% CI +0.030, +0.108; *P* < .001; *I*^2^ = 1.6%; Supplementary Table S10).

### External Validation in OASIS-3

OASIS-3 (*N* = 428; *N* = 425 in M3 Cox) supported RAPS’s prognostic value (HR = 1.35 [1.05, 1.74], *P* = .018); RAPS yielded AUC = 0.864 vs CL 0.833 at slope > 1.0/year (bootstrap *P* = .569; Supplementary Figure S10; Supplementary Table S8).

The intermediate-range (CL 20–60) effect observed in ADNI was partially replicated in OASIS-3 (*n* = 89; Cohen’s *d* = 0.560, *P* = .010; Supplementary Figure S7).

The CL > 0 sensitivity model included 715 participants and selected seven regions. In the same external participants, AUCs for the original nine-region RAPS and the CL > 0 model were 0.803 and 0.794 in NACC SCAN and 0.864 and 0.855 in OASIS-3. Paired differences for the CL > 0 model minus original RAPS were −0.009 (*P* = .067) and −0.010 (*P* = .147), respectively. When both models were retrained using the same corrected inputs and slope-estimation procedure, AUC differences for CL > 0 minus CL ≥ 20 were +0.005 (*P* = .107) in NACC SCAN and −0.009 (*P* = .037) in OASIS-3. These exploratory comparisons were unadjusted for multiplicity and conditional on the fitted models; nonsignificant differences do not establish equivalence. The primary nine-region RAPS was unchanged (Supplementary Table S11).

In the NACC SCAN CL ≥ 20 subset (539 participants; 110 Cox events), the M3 RAPS HR was 1.19 [0.97–1.46] (*P* = .097); AUCs were 0.725 for RAPS and 0.705 for CL (*P* = .559; Supplementary Table S12).

The ancillary tau analysis included 70 ADNI participants and 217 post-index CDR observations. The RAPS coefficient was 0.073 CDR-SB points/year per original discovery-cohort SD before tau adjustment (95% CI −0.051 to 0.196; *P* = .244) and 0.057 after adjustment (95% CI −0.063 to 0.177; *P* = .345). The tau coefficient was 0.133 per subset SD (*P* = .021). RAPS was not statistically significant either before or after tau adjustment in this small selected subset (Supplementary Table S13).

After excluding the 13 participants with one retained CDR-SB observation, the original nine-region set was refitted using corrected inputs and slopes re-estimated in the remaining 420 participants, yielding AUCs of 0.799 in NACC SCAN and 0.876 in OASIS-3, compared with 0.803 and 0.864 for the original RAPS. The NACC M3 HR was 1.19 (95% CI 0.99–1.44; *P* = .061). Repeating region selection in the corrected 420-participant dataset yielded five regions under the original seed and selection criterion, with external AUCs of 0.777 in NACC SCAN and 0.886 in OASIS-3. CL-adjusted associations after region reselection varied across cohorts. The original model was retained for the primary analyses; coefficient, region-reselection and pooled comparisons are reported in Supplementary Table S14.

## Discussion

### Principal findings

This study shows that regional amyloid PET patterns contain prognostic information beyond global Centiloid. Using ElasticNet with bootstrap stability assessment in the ADNI discovery cohort (*N* = 433), we derived RAPS, a compact 9-region score from 82 FreeSurfer ROIs. In ADNI post-selection evaluation, RAPS correlated more strongly with CDR-SB slope than Centiloid (*r* = 0.545 vs 0.314; Steiger *P* < .001) and discriminated rapid decliners more accurately (AUC 0.813 vs 0.708 at >1.0/yr), with positive net reclassification and integrated discrimination improvement estimates in exploratory descriptive analyses, though absolute magnitudes were modest. In the principal external validation—the NACC SCAN multi-tracer cohort (*N* = 1531, four tracers)—RAPS remained an independent predictor of decline alongside Centiloid (M3 Cox HR = 1.20 per SD, *P* = .043) and showed numerically higher discrimination of rapid decliners (AUC 0.803 vs 0.750). RAPS remained associated with clinical progression after adjustment for Centiloid in both external cohorts. AUC differences favored RAPS numerically, but were not statistically significant within either external cohort (NACC SCAN, *P* = .055; OASIS-3, *P* = .569). Across the three independent cohorts, random-effects meta-analysis favored RAPS alone over Centiloid alone (pooled ΔAUC = +0.069; 95% CI +0.030, +0.108; *I*^2^ = 1.6%). In the ADNI development cohort, RAPS was associated with decline and showed higher discrimination than Centiloid in both enrollment-defined CN and MCI strata.

### Beyond Centiloid

In the NACC SCAN cohort (*N* = 1531, four tracers), RAPS and Centiloid each retained independent prognostic value in multivariable Cox models (M3; Figure 4C). Across the four dataset-specific M3 analyses (including the overlapping ADNI FBB subgroup), RAPS hazard ratios ranged from 1.20 to 1.61 per SD. Cross-cohort meta-analysis favored RAPS alone over Centiloid alone (Figure 4D); this comparison does not estimate the AUC gain from adding RAPS to a Centiloid model. This is consistent with the established temporal lag between amyloid pathology and cognitive change.

Prognostic stratification of amyloid-positive individuals is an unmet clinical need in the era of disease-modifying therapy. The CL 20–60 ‘intermediate-range’ effect (ADNI Cohen’s *d* = 0.947) was partially replicated in OASIS-3. Across the overall cohorts, higher discrimination of RAPS alone relative to Centiloid alone was supported by pooled cross-cohort analysis (Figure 4D), supporting prognostic stratification over multi-year follow-up. The CL 20–60 prognostic stratum is distinct from the diagnostic ‘gray zone’ of 11–26 CL reported around data-driven and visual positivity thresholds of 18 and 27 CL.^46^ Our subgroup analysis examined prognostic differences within the amyloid-positive range. Interpretation should be guided by the clinical context.^47^ Whether RAPS can further identify at-risk individuals below conventional positivity thresholds is an important future direction.

### Interpretation

Among the 9 regions, the left hippocampus had the largest absolute coefficient (β = −0.169) with 100% bootstrap selection. Higher hippocampal SUVR was associated with slower decline; hippocampal SUVR alone had a lower AUC than the nine-region RAPS (0.666 vs 0.813), supporting the combined regional score over hippocampal SUVR alone. The inverse association between hippocampal SUVR and CDR-SB slope persisted after hippocampal volume adjustment (partial *r* = −0.231, *P* < .001; Supplementary Methods) and was accompanied by a lower hippocampal-to-cortical SUVR ratio in rapid than slow decliners (0.72 vs 0.84); this observational contrast does not identify its biological cause. Positive fusiform and transverse temporal coefficients and negative entorhinal and pallidal coefficients describe a multivariable prediction pattern and do not establish region-specific harmful or protective effects. ElasticNet coefficients represent predictive weights, not direct mechanisms; without PVE-correction,^29^ hippocampal SUVR may partly reflect atrophy-related signal loss. RAPS should be viewed as a prognostic composite, not a mechanistic amyloid map; replication with PVE-corrected PET is needed. Whereas the Centiloid target region is a large, fixed cortical volume designed to summarize global burden, RAPS uses a small set of FreeSurfer cortical and subcortical regions with signed, data-driven weights, so the two quantify different aspects of the amyloid signal. Whether the left hippocampal contribution reflects atrophy rather than amyloid is examined in Supplementary Methods; partial-volume effects were not corrected, and incorporating MR-based partial-volume correction or atrophy adjustment is a natural extension of RAPS. The regional score may partly reflect correlates of tau pathology or neurodegeneration. The limited tau-adjusted analysis did not establish a tau-independent RAPS association, and these data do not support a mechanistic claim that regional amyloid topology itself drives decline.

Model development focused on amyloid-positive CN/MCI participants, whereas external validation included broader cohorts (NACC included dementia cases). CDR-SB slope captured the dynamic trajectory of decline, more clinically relevant than single-visit status. Regional approaches are gaining traction across molecular imaging: standardized tau PET scales and in vivo Braak staging capture the spatial distribution of tau and predict cognitive decline.^48–49^ RAPS applies an analogous spatial logic to amyloid, deriving a data-driven regional weighting optimized for prognosis rather than for cross-tracer harmonization. Amyloid accumulation follows a sigmoidal trajectory that plateaus at high burden,^50–51^ which may limit the prognostic resolution of global amyloid metrics once positivity is established; downstream markers such as FDG-PET add complementary predictive information.^52^ RAPS summarizes regional amyloid PET measurements for prognosis; their association with decline may also reflect other disease processes.

Disease stage and amyloid burden also differ across cohorts. Mean CL was 74.1 in ADNI, 23.6 in NACC SCAN and 25.4 in OASIS-3; the external cohorts had larger proportions with baseline CDR-SB = 0 (Table 1). These differences in case mix and outcome range may contribute to attenuated external effects, alongside tracer and processing differences. The CL 20–60 subgroup defines amyloid burden rather than cognitive severity. In the NACC CL ≥ 20 subset, the M3 HR was similar to that in the full cohort, but its confidence interval included 1 and the AUC difference was not significant.

### Clinical implications

Mattsson et al.^16^ developed a regional amyloid PET staging approach, but systematic data-driven multi-cohort validation has been limited. RAPS extends this through ADNI training and multi-cohort, multi-tracer external validation, and may complement Centiloid for prognostic stratification, pending prospective validation. RAPS was optimized for CDR-SB slope whereas Centiloid quantifies total amyloid burden, an asymmetry relevant to comparative performance. In settings where Centiloid quantification is available, RAPS can use regional measurements from the same amyloid PET scan. CL is required to identify a CL-defined subgroup such as CL 20–60, but is not an input to the RAPS formula. Supervised optimization against the outcome and post-selection evaluation can favor RAPS in ADNI; the external CL-adjusted associations and the nonsignificant external AUC comparisons should therefore be distinguished.

The 9-ROI model simplifies RAPS computation to a subset of standard FreeSurfer regions, facilitating integration into existing pipelines. Fixed-weight performance was evaluated without retraining in the multi-tracer NACC SCAN cohort, with directionally consistent tracer-stratified associations, but formal equivalence across tracers was not established; the ADNI florbetaben subgroup was small and largely overlapped the discovery cohort, so validation in larger, independent florbetaben cohorts is needed. Regionally weighted quantification may also complement visual reads, which can be challenging in cases with focal or low-level deposition; quantitative support improves reader agreement and confidence in such cases.^53^

### Limitations

Limitations include: (i) potential informative dropout bias; (ii) all cohorts are predominantly white, and OASIS-3 differed from ADNI in composition and reference regions; (iii) post-selection CV showed fold-level variability (Supplementary Figure S2);^54–56^ (iv) PVE was not corrected;^29^ (v) external analyses used the ADNI-fitted StandardScaler; (vi) results may depend on the parcellation^26^ and hyperparameter grid; (vii) the NACC analytic cohort included dementia cases (*n* = 171) whereas ADNI was restricted to CN/MCI; (viii) RAPS was derived from two [^18^F] tracers (florbetapir and florbetaben) whose regional SUVRs were pooled without tracer harmonization or tracer adjustment, and although tracer-stratified associations in the multi-tracer NACC SCAN cohort (florbetapir, florbetaben, PiB, and NAV4694) were directionally consistent, regional uptake patterns can differ across tracers, formal tracer equivalence was not established, and residual tracer-specific bias cannot be excluded; (ix) the nine RAPS regions were selected on the full discovery sample, so the internal out-of-fold performance is a post-selection cross-validation that may be optimistic for the feature-selection step, with generalizability supported primarily by the fixed-weight external cohorts; and (x) the ADNI [^18^F]florbetaben subgroup largely overlapped the discovery cohort (a small-sample sensitivity analysis rather than independent validation), and participant overlap between NACC/OASIS-3 and ADNI cannot be fully excluded owing to differing identifier systems. Caudate values from different PET scans in the original inputs were examined in corrected-input sensitivity analyses, alongside analyses excluding the 13 single-observation participants (Supplementary Tables S11 and S14). Refitting the original nine regions produced small changes in external AUCs, whereas de novo selection yielded different region sets. The predictive performance of the fixed score should therefore be distinguished from the stability of its selected region set. RAPS was developed in a single ADNI cohort. Larger, multicohort development studies could further assess the stability of regional selection, with independent validation of any updated model. Future directions include validation in Japanese (JADNI) cohorts, tau PET integration,^57–58^ and voxel-level analysis.

### Conclusions

RAPS, an amyloid PET-derived regional prognostic score, may complement rather than replace Centiloid for prognostic stratification. Cross-cohort synthesis, including the post-selection ADNI development-cohort estimate, favored RAPS alone over Centiloid alone (pooled ΔAUC = +0.069; *I*^2^ = 1.6%), warranting prospective validation.

## Acknowledgements

Data collection and sharing for ADNI is funded by the National Institute on Aging (National Institutes of Health Grant U19 AG024904). The grantee organization is the Northern California Institute for Research and Education. In the past, ADNI has also received funding from the National Institute of Biomedical Imaging and Bioengineering, the Canadian Institutes of Health Research, and private sector contributions through the Foundation for the National Institutes of Health (FNIH) including generous contributions from the following: AbbVie, Alzheimer’s Association; Alzheimer’s Drug Discovery Foundation; Araclon Biotech; BioClinica, Inc.; Biogen; Bristol-Myers Squibb Company; CereSpir, Inc.; Cogstate; Eisai Inc.; Elan Pharmaceuticals, Inc.; Eli Lilly and Company; EuroImmun; F. Hoffmann-La Roche Ltd and its affiliated company Genentech, Inc.; Fujirebio; GE Healthcare; IXICO Ltd.; Janssen Alzheimer Immunotherapy Research & Development, LLC.; Johnson & Johnson Pharmaceutical Research & Development LLC.; Lumosity; Lundbeck; Merck & Co., Inc.; Meso Scale Diagnostics, LLC.; NeuroRx Research; Neurotrack Technologies; Novartis Pharmaceuticals Corporation; Pfizer Inc.; Piramal Imaging; Servier; Takeda Pharmaceutical Company; and Transition Therapeutics.

Data were provided in part by OASIS-3: Longitudinal Multimodal Neuroimaging, Clinical, and Cognitive Dataset for Normal Aging and Alzheimer’s Disease. Principal Investigators: T. Benzinger, D. Marcus, and J. Morris. OASIS-3 is supported by NIH P30 AG066444, P50 AG00561, P30 NS09857781, P01 AG026276, P01 AG003991, R01 AG043434, UL1 TR000448, and R01 EB009352. AV-45 doses were provided by Avid Radiopharmaceuticals, a wholly owned subsidiary of Eli Lilly.

Data were also provided in part by the National Alzheimer’s Coordinating Center (NACC) and its Standardized Centralized Alzheimer’s & Related Dementias Neuroimaging (SCAN) initiative. The NACC database is funded by NIA/NIH Grant U24 AG072122. NACC data are contributed by the NIA-funded ADRCs: P30 AG062429 (PI James Brewer, MD, PhD), P30 AG066468 (PI Oscar Lopez, MD), P30 AG062421 (PI Teresa Gomez-Isla, MD), P30 AG066509 (PI Thomas Grabowski, MD), P30 AG066514 (PI Mary Sano, PhD), P30 AG066530 (PI Helena Chui, MD, Arthur Toga, PhD), P30 AG066507 (PI Marilyn Albert, PhD), P30 AG066444 (PI David Holtzman, MD), P30 AG066518 (PIs Lisa Silbert, MD, Kevin Duff, PhD), P30 AG066512 (PI Thomas Wisniewski, MD), P30 AG066462 (PI Scott Small, MD), P30 AG072979 (PI David Wolk, MD), P30 AG072972 (PIs Charles DeCarli, MD, Rachel Whitmer, PhD), P30 AG072976 (PI Andrew Saykin, PsyD), P30 AG072975 (PI Julie Schneider, MD, MS), P30 AG072978 (PI Ann McKee, MD), P30 AG072977 (PI Robert Vassar, PhD), P30 AG066519 (PI Joshua Grill, PhD), P30 AG062677 (PIs Brad Boeve, MD, Ronald Petersen, MD, PhD), P30 AG079280 (PI Jessica Langbaum, PhD), P30 AG062422 (PI Gil Rabinovici, MD), P30 AG066511 (PI Allan Levey, MD, PhD), P30 AG072946 (PI Linda Van Eldik, PhD), P30 AG062715 (PI Sanjay Asthana, MD, FRCP), P30 AG072973 (PI Russell Swerdlow, MD), P30 AG066506 (PIs Glenn Smith, PhD, ABPP, David Lowenstein, PhD, Ranjan Duara, MD), P30 AG066508 (PIs Stephen Strittmatter, MD, PhD, Christopher Van Dyck, MD), P30 AG066515 (PI Victor Henderson, MD, MS), P30 AG072947 (PI Suzanne Craft, PhD), P30 AG072931 (PI Henry Paulson, MD, PhD), P30 AG066546 (PIs Sudha Seshadri, MD, Gladys Maestre, MD, PhD), P30 AG086401 (PI Erik Roberson, MD, PhD), P30 AG086404 (PI Gary Rosenberg, MD), P30 AG086403 (PI Angela Jefferson, PhD), P30 AG072958 (PIs Heather Whitson, MD, Gwenn Garden, MD, PhD), P30 AG072959 (PI Jagan Pillai, MD, PhD), P30 AG092752 (Ihab Hajjar, MD, MS). SCAN is a multi-institutional project that was funded as a U24 grant (AG067418) by the National Institute on Aging in May 2020. Data collected by SCAN and shared by NACC are contributed by the NIA-funded ADRCs as follows: Arizona Alzheimer’s Center - P30 AG072980 (PI: Eric Reiman, MD); R01 AG069453 (PI: Eric Reiman (contact), MD); P30 AG019610 (PI: Eric Reiman, MD); and the State of Arizona which provided additional funding supporting our center; Boston University - P30 AG013846 (PI Neil Kowall MD); Cleveland ADRC - P30 AG062428 (James Leverenz, MD); Cleveland Clinic, Las Vegas - P20AG068053; Columbia - P50 AG008702 (PI Scott Small MD); Duke/UNC ADRC - P30 AG072958; Emory University - P30AG066511 (PI Levey Allan, MD, PhD); Indiana University - R01 AG19771 (PI Andrew Saykin, PsyD); P30 AG10133 (PI Andrew Saykin, PsyD); P30 AG072976 (PI Andrew Saykin, PsyD); R01 AG061788 (PI Shannon Risacher, PhD); R01 AG053993 (PI Yu-Chien Wu, MD, PhD); U01 AG057195 (PI Liana Apostolova, MD); U19 AG063911 (PI Bradley Boeve, MD); and the Indiana University Department of Radiology and Imaging Sciences; Johns Hopkins - P30 AG066507 (PI Marilyn Albert, Phd.); Mayo Clinic - P50 AG016574 (PI Ronald Petersen MD PhD); Mount Sinai - P30 AG066514 (PI Mary Sano, PhD); R01 AG054110 (PI Trey Hedden, PhD); R01 AG053509 (PI Trey Hedden, PhD); New York University - P30AG066512-01S2 (PI Thomas Wisniewski, MD); R01AG056031 (PI Ricardo Osorio, MD); R01AG056531 (PIs Ricardo Osorio, MD; Girardin Jean-Louis, PhD); Northwestern University - P30 AG013854 (PI Robert Vassar PhD); R01 AG045571 (PI Emily Rogalski, PhD); R56 AG045571, (PI Emily Rogalski, PhD); R01 AG067781, (PI Emily Rogalski, PhD); U19 AG073153, (PI Emily Rogalski, PhD); R01 DC008552, (M.-Marsel Mesulam, MD); R01 AG077444, (PIs M.-Marsel Mesulam, MD, Emily Rogalski, PhD); R01 NS075075 (PI Emily Rogalski, PhD); R01 AG056258 (PI Emily Rogalski, PhD); Oregon Health and Science University - P30 AG066518 (PIs Lisa Silbert, MD, Kevin Duff, PhD); R56 AG074321 (PI Jeffrey Kaye, MD); Rush University - P30 AG010161 (PI David Bennett MD); Stanford - P30AG066515; P50 AG047366 (PI Victor Henderson MD MS); University of Alabama, Birmingham - P20; University of California, Davis - P30 AG10129 (PI Charles DeCarli, MD); P30 AG072972 (PI Charles DeCarli, MD); University of California, Irvine - P50 AG016573 (PI Frank LaFerla PhD); University of California, San Diego - P30AG062429 (PI James Brewer, MD, PhD); University of California, San Francisco - P30 AG062422 (Rabinovici, Gil D., MD); University of Kansas - P30 AG035982 (Russell Swerdlow, MD); University of Kentucky - P30 AG028283-15S1 (PIs Linda Van Eldik, PhD and Brian Gold, PhD); University of Michigan ADRC - P30AG053760 (PI Henry Paulson, MD, PhD) P30AG072931 (PI Henry Paulson, MD, PhD) Cure Alzheimer’s Fund 200775 - (PI Henry Paulson, MD, PhD) U19 NS120384 (PI Charles DeCarli, MD, University of Michigan Site PI Henry Paulson, MD, PhD) R01 AG068338 (MPI Bruno Giordani, PhD, Carol Persad, PhD, Yi Murphey, PhD) S10OD026738-01 (PI Douglas Noll, PhD) R01 AG058724 (PI Benjamin Hampstead, PhD) R35 AG072262 (PI Benjamin Hampstead, PhD) W81XWH2110743 (PI Benjamin Hampstead, PhD) R01 AG073235 (PI Nancy Chiaravalloti, University of Michigan Site PI Benjamin Hampstead, PhD) 1I01RX001534 (PI Benjamin Hampstead, PhD) IRX001381 (PI Benjamin Hampstead, PhD); University of New Mexico - P20 AG068077 (Gary Rosenberg, MD); University of Pennsylvania - State of PA project 2019NF4100087335 (PI David Wolk, MD); Rooney Family Research Fund (PI David Wolk, MD); R01 AG055005 (PI David Wolk, MD); University of Pittsburgh - P50 AG005133 (PI Oscar Lopez MD); University of Southern California - P50 AG005142 (PI Helena Chui MD); University of Washington - P50 AG005136 (PI Thomas Grabowski MD); University of Wisconsin - P50 AG033514 (PI Sanjay Asthana MD FRCP); Vanderbilt University - P20 AG068082; Wake Forest - P30AG072947 (PI Suzanne Craft, PhD); Washington University, St. Louis - P01 AG03991 (PI John Morris MD); P01 AG026276 (PI John Morris MD); P20 MH071616 (PI Dan Marcus); P30 AG066444 (PI John Morris MD); P30 NS098577 (PI Dan Marcus); R01 AG021910 (PI Randy Buckner); R01 AG043434 (PI Catherine Roe); R01 EB009352 (PI Dan Marcus); UL1 TR000448 (PI Brad Evanoff); U24 RR021382 (PI Bruce Rosen); Avid Radiopharmaceuticals / Eli Lilly; Yale - P50 AG047270 (PI Stephen Strittmatter MD PhD); R01AG052560 (MPI: Christopher van Dyck, MD; Richard Carson, PhD); R01AG062276 (PI: Christopher van Dyck, MD); 1Florida - P30AG066506-03 (PI Glenn Smith, PhD); P50 AG047266 (PI Todd Golde MD PhD).

During the preparation and revision of this manuscript, the authors used Claude (Anthropic), ChatGPT and Codex (OpenAI) to assist with analysis-code development, data-provenance and numerical-consistency checks, reference checking, and manuscript, figure and table preparation. AI-assisted outputs were treated as drafts requiring verification against source data and analysis code. The authors are responsible for the scientific decisions and the accuracy and integrity of the final content.

## Author Contributions

T.H. conceived and designed the study, developed the analysis pipeline, curated and processed the data, performed all statistical analyses, generated the figures and tables, acquired funding, and drafted the manuscript. W.A. contributed to the conceptualization of the study, acquired funding, and contributed to manuscript revision. T.K. contributed to the conceptualization of the study and the clinical framing of the research question, provided clinical expertise, and contributed to manuscript revision. All authors read and approved the final manuscript.

## Statements and Declarations

### Ethical Considerations

This study was approved by the Institutional Review Board of Juntendo University (approval numbers: E23-0461, E25-0488) and was conducted in accordance with the Declaration of Helsinki. All participants in ADNI, OASIS-3, and NACC SCAN provided written informed consent at their respective study sites.

### Consent to Participate

All participants (or their authorized representatives) provided written informed consent at each contributing study site under protocols approved by the respective local institutional review boards. The present work is a secondary analysis of de-identified data and required no additional consent.

### Consent for Publication

Not applicable.

### Funding

This work was supported in part by Japan Society for the Promotion of Science (JSPS) KAKENHI (Grant Number JP24K18745 to T.H.) and the Japan Agency for Medical Research and Development (AMED), Basic Research Program for Drug Discovery Promotion (GAPFREE; Grant Number JP25ak0101236 to W.A.).

### Declaration of Conflicting Interests

The author(s) declared no potential conflicts of interest with respect to the research, authorship, and/or publication of this article.

### Data Availability Statement

The datasets supporting the conclusions of this article are publicly available from third-party repositories. ADNI data are available from the Alzheimer’s Disease Neuroimaging Initiative (https://adni.loni.usc.edu/) upon approval of a Data Use Agreement and review by the ADNI Data Sharing and Publications Committee. OASIS-3 data are available from https://www.oasis-brains.org/ (NITRC server: https://nitrc.org/ir/) upon application. NACC SCAN data are available from the National Alzheimer’s Coordinating Center (https://naccdata.org/) upon Data Request approval. Analysis code, the final 9-ROI RAPS model weights, and a turnkey computation script will be made publicly available at https://github.com/t-hirose-lab/raps-regional-amyloid-pet-score upon publication. Reviewer access can be provided upon request.

## Supplementary information

The Supplementary Information contains 14 Tables (S1–S14) and 10 Figures (S1–S10): 82-ROI bootstrap (S1), reproducibility parameters (S2), 9-ROI RAPS weights (S3), cross-cohort distributional summary (S4), subgroup analyses (S5), *APOE* × RAPS interaction (S6), Cox PH per cohort (S7), AUC per cohort/threshold (S8), NACC tracer-stratified correlations (S9), meta-analysis ΔAUC details (S10), CL-range sensitivity retraining (S11), NACC CL ≥ 20 sensitivity (S12), the ancillary ADNI tau analysis (S13), and sensitivity analyses excluding single-observation development participants (S14). Figures S1–S4: OLS vs BLUP, post-selection CV, bootstrap CIs, 9-ROI vs 82-ROI comparison; S5: subgroup analysis and clinical utility; S6: ADNI survival; S7: CL 20–60 exploratory; S8: NACC tracer forest; S9: reclassification (NRI/IDI); S10: OASIS-3 ROC.

## Notes

### Competing Interest Statement

The authors have declared no competing interest.

### Author Declarations

The Institutional Review Board of Juntendo University gave ethical approval for this work (approval numbers E23-0461 and E25-0488). This study is a secondary analysis of de-identified data; all participants provided written informed consent at the original study sites (ADNI, NACC, and OASIS-3) under protocols approved by the respective institutional review boards.

### Summary of Updates

The manuscript, figures, tables, and supplementary materials have been updated to reflect the version accepted for publication in the Journal of Alzheimer's Disease. Changes include clarification of cohort composition and participant overlap, updated results, and additional sensitivity analyses.

