## Supplementary_Information for "A data-driven regional amyloid PET score predicts cognitive decline beyond Centiloid"

### Supplementary Methods

#### CDR-SB longitudinal slope estimation

In ADNI, per-subject CDR-SB slopes were estimated via linear mixed-effects (LME) models with random intercept and slope, using the lme4 R package (v1.1.38 for the corrected evaluation). Sensitivity analysis comparing LME-derived BLUP slopes with simple ordinary least-squares (OLS) slopes is presented in Supplementary Figure S1. The revision used the recorded CDR assessment dates and removed identical repeated observations, retaining the original clinical-series index and eligibility rule (2,943 observations from 579 participants). These participant-level slopes do not necessarily represent decline exclusively after the PET scan used to compute RAPS.

#### External cohort RAPS application

The 9-ROI RAPS weights (Supplementary Table S3) were applied without retraining to the overlapping ADNI [$^{18}$F]florbetaben subgroup (sensitivity analysis) and the two external validation cohorts (NACC SCAN, OASIS-3), using the fitted StandardScaler from the ADNI discovery cohort in all three datasets. Regional SUVRs were used as supplied by each cohort: whole-cerebellum-referenced SUVRs in ADNI and NACC SCAN, and cerebellar-cortex-referenced PUP regional SUVRs in OASIS-3 (OASIS-3 Imaging Data Dictionary v2.0, p. 24). No additional re-referencing was applied. For OASIS-3, Centiloid was matched to the scan supplying the nine regional inputs. For three participants without a same-scan entry in the dedicated Centiloid table, non-PVC mean cortical SUVR from that PUP record was converted using CL = 163.6 × SUVR − 181.0 (OASIS-3 Imaging Data Dictionary v2.0, Table 14, Eq. 5). For NACC SCAN, participant-level slopes were estimated by ordinary least-squares regression of CDR-SB on elapsed years using all available valid clinical observations (CDR-SB 0–18), including observations before PET; eligibility required at least two observations spanning at least six months. The retained slopes, observation counts and observation spans were reproduced for all 1,531 participants. The baseline CDR-SB covariate was the valid assessment closest to PET, whereas the Cox event threshold used the first assessment on or after PET. For OASIS-3, slopes were estimated jointly with a random-intercept/random-slope linear mixed-effects model fitted by restricted maximum likelihood (REML), using CDR observations on or after the earliest AV45 PET scan. Participants required at least two observations and a latest observation at least two years after PET. The LME branch reproduced all 429 stored slopes at their six-decimal precision; no OLS fallback was needed in this verification. Table 1 retained the CDR-SB assessment closest to PET, whereas the Cox baseline covariate and event threshold used the first assessment on or after PET. These original external-cohort slopes were retained in the fixed-score evaluation.

**NACC Cox proportional-hazards formulation**

For NACC SCAN, visit-level CDR-SB data were obtained from the NACC investigator file (investigator_nacc73.csv) and merged with the SCAN PET dataset. Cox proportional-hazards models used proper time-to-event analysis: event = first post-PET visit with CDR-SB ≥ baseline + 1.0; censoring at last visit; time = years from PET to event or censoring (178 events / 1531 subjects). The ROC analysis (Figure 4B) uses a separate slope-based binary outcome (CDR-SB slope > 1.0/year; 117 events) for direct comparison with ADNI and OASIS-3 figures. Education code 99 was treated as missing; M3 included 1,529 participants and 178 events. The original PET time origin and baseline CDR covariate were retained. APOE ε4 allele counts were derived from the official NACCAPOE coding (codes 1, 3 and 6: zero; codes 2 and 5: one; code 4: two) and checked against NACCNE4S in the source data.

**Hippocampal volume adjustment**

To assess whether the left hippocampus negative coefficient reflects atrophy-related signal loss rather than amyloid burden, we computed the partial correlation between left hippocampal SUVR and unadjusted CDR-SB slope after residualizing both variables on left hippocampal volume ($mm^3$, from FreeSurfer aseg subcortical segmentation; no intracranial-volume normalization). Partial $r$ was computed by ordinary least-squares residualization in the ADNI discovery cohort ($N$ = 433). Higher hippocampal SUVR remained associated with slower CDR-SB decline after this adjustment (partial $r = -0.231$, $P < .001$), although this partial correlation does not exclude partial-volume or other atrophy-related effects.

**Bootstrap feature-selection procedure**

ROI selection stability was assessed via 1,000 bootstrap iterations (resampling with replacement from the ADNI discovery cohort). In each iteration, an

ElasticNetCV model (L1 ratio grid {0.1, 0.3, 0.5, 0.7, 0.9}, 5-fold inner cross-validation, random_state = 42) was refit de novo on the resampled data, and the frequency with which each of the 82 ROIs received a non-zero coefficient was recorded as its bootstrap selection frequency.

**ROI selection and final refit sequence**

The 9-ROI RAPS was defined in four sequential steps: (1) bootstrap selection frequency was computed for all 82 ROIs as described above; (2) a single ElasticNetCV model was separately fit on the complete discovery sample (same hyperparameter grid) to obtain full-data coefficients; (3) ROIs satisfying both the intersection criterion (bootstrap selection frequency ≥ 80% and non-zero full-data coefficient) were retained, yielding 9 ROIs; (4) the 9-ROI model was internally validated via 5×5 post-selection cross-validation (Supplementary Figure S2), and a final model was refit on the complete discovery sample using these 9 ROIs to obtain the fixed coefficients (Supplementary Table S3) applied to all external cohorts. During the revision audit, the nine regional inputs used by the locked RAPS model and the corresponding Centiloid value were matched to a single raw PET record for each of the 433 ADNI discovery participants. The historical 82-region input matrix contained caudate values from a different scan in 283 participants. Substituting caudate values from the matched scan left predictions from the locked nine-region model unchanged. The nine regions, fitted coefficients, input scaler and saved ADNI post-selection out-of-fold scores were retained. The primary model was not refitted; additional feature selection and refitting using corrected inputs were restricted to the CL-range and single-observation-exclusion sensitivity analyses described below. The original nine-region internal cross-validation and CL-stratified coefficient analyses were also retained without refitting (Supplementary Figures S2–S4 and S5B); these describe model development using the original outcomes, whereas the revised correlation, discrimination and survival results evaluate the fixed scores using corrected data.

**External cohort standardization**

Regional SUVRs for the fixed 9-ROI RAPS were standardized using the StandardScaler fitted on the ADNI discovery cohort (mean and SD locked at derivation; no re-fitting in external cohorts). For comparative effect-size analyses (e.g., Cohen's *d*, median-split survival analyses), the resulting RAPS predictions were subsequently standardized within each cohort.

**Meta-analysis variance estimation**

For the pooled ΔAUC (RAPS alone − Centiloid alone) analysis, the standard error of each cohort-level ΔAUC estimate was obtained empirically via paired bootstrap resampling (1,000 iterations, resampling participants with replacement and recomputing both AUCs in each iteration); no between-score correlation was assumed a priori. The corresponding $z$ statistics and $P$ values were computed directly from this bootstrap standard error ($z = \Delta AUC / SE$; two-sided $P$ from the standard normal distribution); the analytic DeLong covariance formulation was not used, and these tests are accordingly referred to as paired bootstrap tests elsewhere in the manuscript. For the pooled hazard ratio (M3) analysis, the variance was taken as the squared standard error of the log hazard ratio from the corresponding Cox model. Cohort-level estimates were pooled using DerSimonian-Laird random-effects meta-analysis, with between-study heterogeneity ($\tau^2$, $I^2$) estimated from Cochran's $Q$. The primary pooled estimates were computed from the three independent cohorts (ADNI discovery, NACC SCAN, OASIS-3); the ADNI [$^{18}$F]florbetaben subgroup, which largely overlaps the discovery cohort (68/71 participants), was excluded from pooling and evaluated separately as a descriptive sensitivity analysis.

**RAPS computation**

The definitive RAPS for a given participant is computed as the sum of each of the 9 fixed regional SUVRs (standardized using the locked ADNI discovery-cohort scaler) multiplied by its corresponding fixed ElasticNet coefficient (Supplementary Table S3).

**Sensitivity retraining across the Centiloid development range**

The original eligibility sequence through the clinical-follow-up step was retained, changing the baseline Centiloid criterion to CL > 0. Of 799 potential participants, 791 met the original post-PET row-count requirement and 715 had complete regional inputs and covariates: all 433 original discovery participants plus 282 with 0 < CL < 20. No new requirement for at least two nonmissing post-PET CDR-SB assessments was introduced. As in the original procedure, participants with one retained CDR-SB observation were included (43/715, including 13/433 original discovery participants); their BLUP slopes depend on the fitted population model rather than directly observed within-person change across multiple retained assessments. PET-preceding assessments were not newly added to slope estimation. All 82 regional SUVRs and CL were verified against the same PET record, including corrected caudate values. The 579 original

LME participants and their 2943 corrected observations were retained, and 230 newly eligible participants were added, yielding an expanded LME pool of 809 participants and 3908 observations. Random-intercept/random-slope REML estimation retained the original clinical-series origin. These re-estimated slopes were used only for sensitivity retraining; the primary saved slopes were not replaced.

**Retraining and external evaluation procedure**

The original statistical procedure was retained: covariate residualization for age, sex, education, APOE ε4 count and baseline CDR-SB; 82 regional inputs; 5×5 cross-validation; L1 ratios 0.1, 0.3, 0.5, 0.7 and 0.9; 1000 participant bootstraps; random seed 42; and intersection of selection frequency ≥80% with nonzero full-data elastic-net coefficients. Selected regions were refitted, with post-selection out-of-fold evaluation; region selection was not nested within the outer folds. Reproduction of the historical nine-region refit matched the saved out-of-fold and full-model scores to numerical precision. The CL > 0 model selected seven regions (Supplementary Table S11). A second sensitivity model was fitted in the 433 participants with CL ≥ 20, using the same corrected 82-region inputs, common expanded-pool slopes and statistical procedure. It selected the same seven regions. Shared participants had identical input and outcome values in the two training datasets; each model used its own training-fitted scaler and coefficients. The ADNI-fitted scaler and coefficients of each sensitivity model were frozen before external evaluation in the same 1531 NACC and 428 OASIS-3 participants used in the main analyses. Current corrected CL values and existing external clinical outcomes were used. Paired participant bootstraps (1000, seed 42) assessed AUC differences for the CL > 0 model minus original RAPS and for CL > 0 minus the retrained CL ≥ 20 model, with normal-approximation *P* values and percentile confidence intervals. These two inferential summaries use different procedures and need not give identical significance decisions. Comparisons were exploratory, unadjusted for multiplicity and conditional on the fitted models, without development-model retraining within the bootstrap. The comparison with the original RAPS also includes differences in corrected development inputs and outcome re-estimation. The comparison between retrained models assesses the training-range change under common processing; neither comparison establishes absence of selection bias.

**NACC amyloid-positive sensitivity**

The fixed nine-region model was evaluated in the 539 participants with baseline CL ≥ 20, without changing PET time origin, baseline covariate definitions, outcome slopes, event rules, APOE coding or education handling. RAPS and CL retained their full-NACC standardization so hazard ratios remain comparable in units. M1–M3 and slope-threshold AUC comparisons followed the existing methods; the subset included 110 Cox events and 84 rapid decliners at slope >1.0/year (Supplementary Table S12). The complete CL-unrestricted cohort remained the primary external analysis.

**Ancillary ADNI tau adjustment**

The nearest available UC Berkeley non-PVC META_TEMPORAL_SUVR flortaucipir measurement was selected for each discovery participant, with ties resolved by the earlier tau scan. The amyloid scan was the original scan underlying the fixed score. PET dates had to differ by no more than 365 days. The ancillary index was the later scan; participants required at least two validated CDR observations on or after that date and a final observation at least two years after it. Seventy complete cases contributed 217 observations. A separate REML random-intercept/random-slope LME estimated CDR-SB slopes in this subset. The fixed discovery-cohort post-selection out-of-fold RAPS score was used, retaining its original standardization. Models adjusted for CL, retained age, sex, education and APOE covariates, and the first CDR-SB after the ancillary index, with and without meta-temporal tau; both models used identical cases. CL and tau were standardized within the subset. Ordinary least-squares coefficients, conventional standard errors and two-sided *t* tests are reported in Supplementary Table S13. Independent linear-algebra calculations reproduced the coefficients and standard errors. This exploratory development-subset analysis is not independent validation and does not establish causal independence from tau; selection optimism and uncertainty in the estimated slopes remain. The primary slopes, model and cohorts were not changed.

**Sensitivity analysis excluding single-observation participants**

The repeat-observation check in the original code counted data rows, including rows with missing CDR-SB, rather than nonmissing assessments. This admitted 13 participants with only one retained CDR-SB observation. We excluded these participants for a sensitivity analysis, leaving 420 participants with at least two distinct retained CDR-SB assessment dates. Corrected scan-level regional inputs and clinical dates were used, and slopes were re-estimated using the 2001 observations from these 420 participants. The original clinical-series origin, covariates and Cox time/event definitions were retained; this did not introduce a new requirement that all assessments follow the PET scan used for RAPS. The nine originally selected regions were refitted using the original covariate adjustment, scaling and 5×5 elastic-net cross-validation procedure (seed 42). The original RAPS coefficients and saved scores were not replaced.

Separately, selection from all 82 regions was repeated with the original 1000-bootstrap, ≥80% frequency and nonzero-coefficient intersection rule. Five blocks of 1000 bootstraps, with starting seeds 42, 1042, 2042, 3042 and 4042, selected 5, 6, 6, 6 and 6 regions. Pooling all 5000 bootstrap counts at ≥80% selected the same five regions as the seed-42 analysis. An exploratory alternative retaining regions reaching ≥80% in any block selected eight regions. These alternatives are reported alongside the fixed-nine-region refit in Supplementary Table S14, without using external performance to select a replacement model.

Comparisons used identical external participants and outcomes. Within ADNI, the original saved scores were additionally evaluated in the same 420 participants against the same re-estimated slopes. AUCs describe CDR-SB slope >1.0/year and compare each score alone with CL alone. M3 adjusted for

CL and the original covariates; AUC inference and DerSimonian–Laird pooling followed the main methods. Three-cohort pooling included ADNI, NACC and OASIS-3; external-only pooling included NACC and OASIS-3. Fixed-region refitting remains conditional on regions originally selected using all 433 participants. The analyses were exploratory, with no prespecified equivalence margin; small performance changes do not establish absence of selection effects. The original nine-region RAPS remains the primary model.

**Software environment**

The corrected evaluations used Python 3.12 with the following key packages: scikit-learn 1.8.0, statsmodels 0.14.6, lifelines 0.30.3, pandas 2.3.3, NumPy 2.3.5, SciPy 1.17.0. For the corrected ADNI evaluation, mixed-effects slopes were estimated in R 4.5.2 with lme4 1.1.38. The retained OASIS-3 slopes were estimated with Python statsmodels. Supplementary Table S2 retains the original model-development settings. The additional 420-participant sensitivity analyses used NumPy 2.4.2.
