## Supplementary_Figures for "A data-driven regional amyloid PET score predicts cognitive decline beyond Centiloid"

### Supplementary Figures S1-S10

Article title: A data-driven regional amyloid PET score predicts cognitive decline beyond Centiloid

Authors: Takumi Hirose, Wado Akamatsu, Tadafumi Kato, for the Alzheimer's Disease Neuroimaging Initiative
Corresponding author: Takumi Hirose, MD, PhD
Center for Genomic and Regenerative Medicine, Juntendo University


Supplementary Figure S1

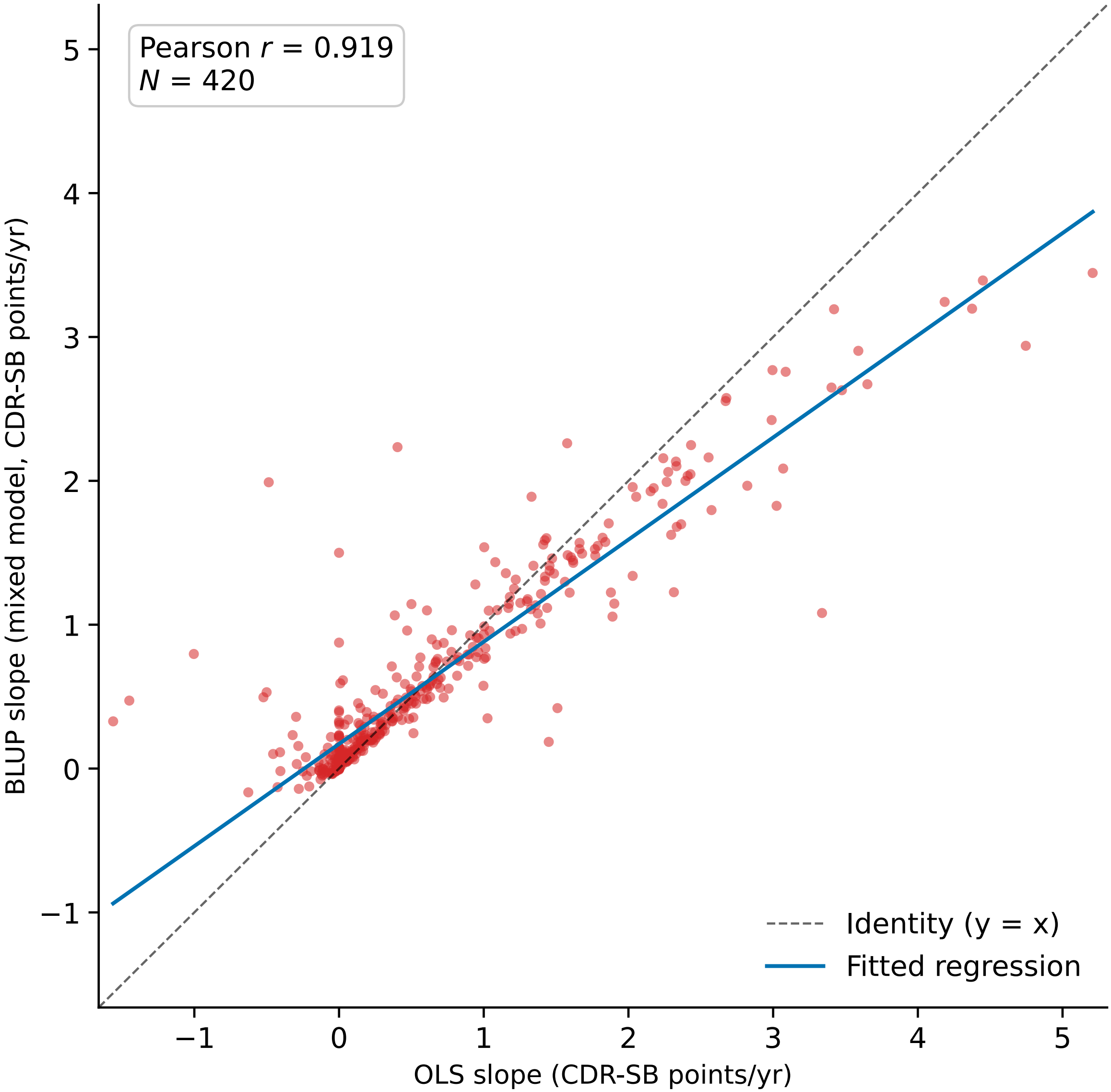

Supplementary Figure S1 OLS vs BLUP Slope Comparison. Scatter plot comparing CDR-SB slopes estimated by ordinary least squares (OLS) and best linear unbiased prediction (BLUP) from the corrected mixed-effects analysis in 420 of the 433 ADNI participants with at least two CDR-SB observations (Pearson *r* = 0.919). The dashed line denotes identity (y = x); the solid blue line is the fitted regression (BLUP = 0.171 + 0.710 × OLS). The departure from identity illustrates shrinkage of the BLUP estimates; high correlation does not imply agreement in magnitude.

Supplementary Figure S2

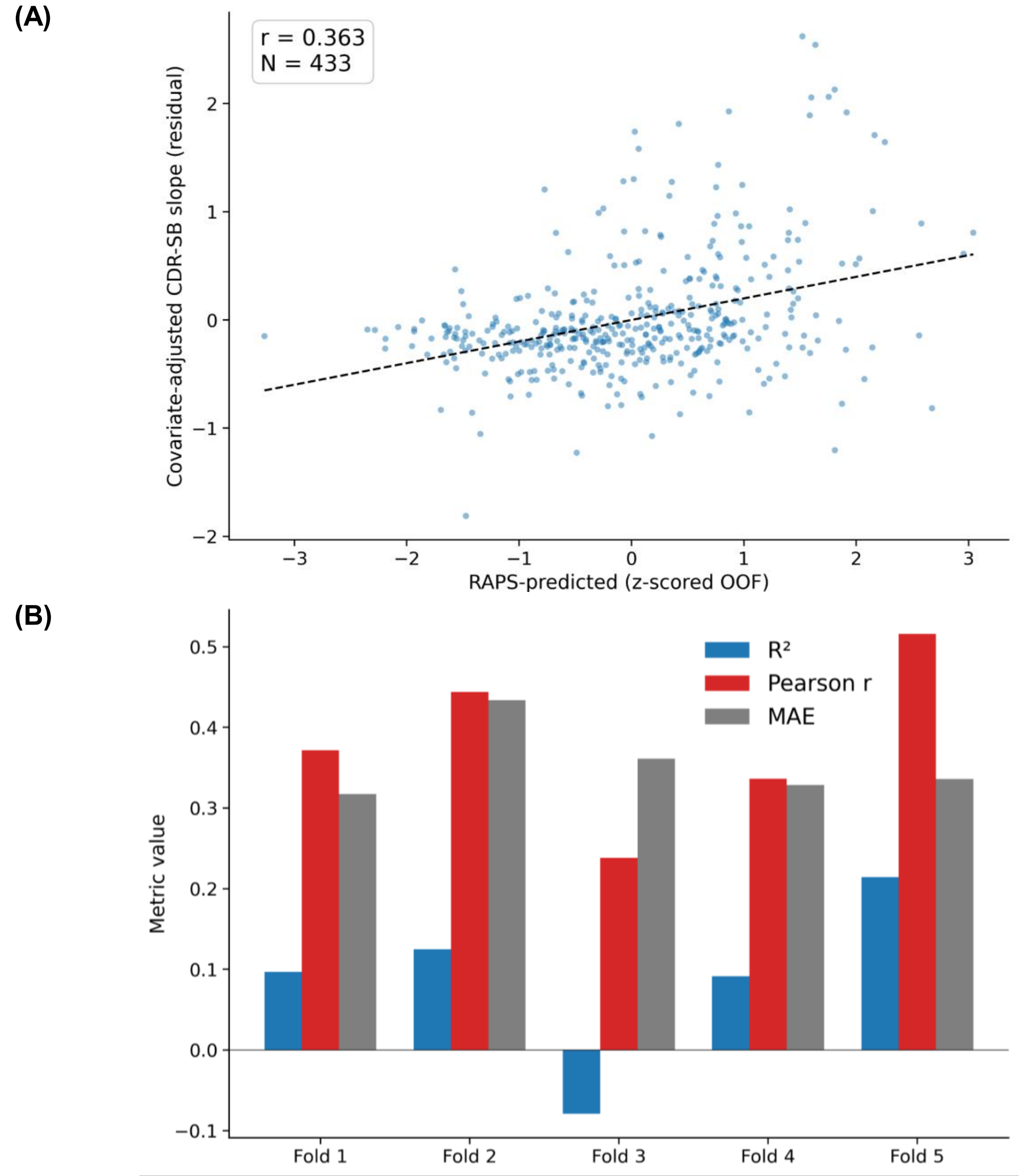

**Supplementary Figure S2 Post-selection Cross-Validation Performance.** (A) Scatter plot of RAPS out-of-fold predictions versus covariate-residualized CDR-SB slope in the ADNI discovery cohort ($N$ = 433; pooled out-of-fold $r$ = 0.363). (B) Per-fold performance metrics ($R^2$, Pearson $r$, MAE) for each of the 5 outer folds in 5×5 post-selection cross-validation, illustrating fold-to-fold variability (fold $R^2$ range: –0.08 to 0.21). Original model-development results, retained without refitting after the data audit.

Supplementary Figure S3

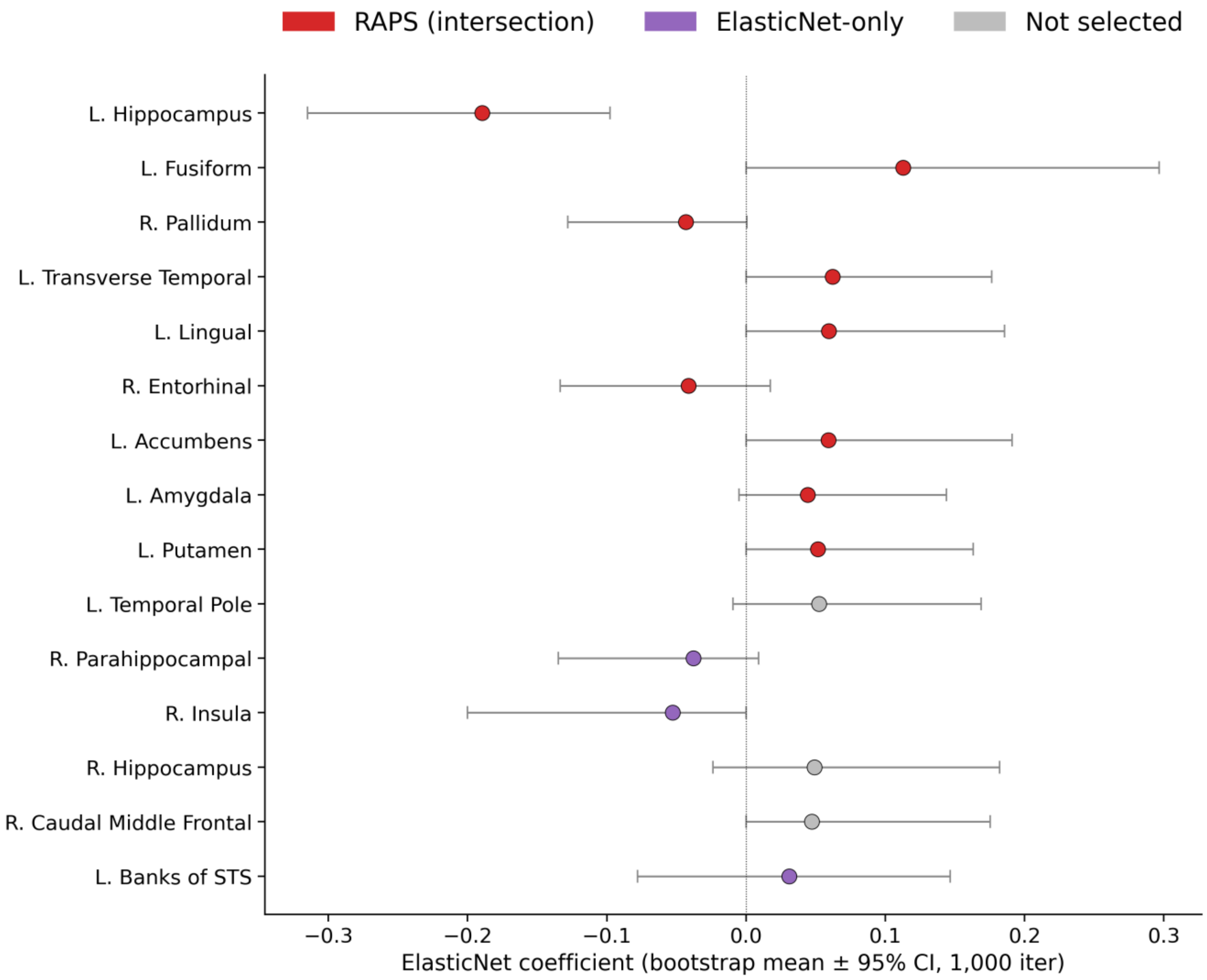

**Supplementary Figure S3 Bootstrap Coefficient 95% Confidence Intervals.**
ElasticNet coefficient estimates with bootstrap means and 95% confidence intervals from 1,000 bootstrap iterations (resampling with replacement, refitting ElasticNet de novo), shown for the top 15 ROIs ranked by bootstrap selection frequency. Red markers ($n$ = 9) denote the intersection ROIs satisfying both criteria (bootstrap selection ≥ 80% and non-zero final ElasticNet coefficient), comprising the final RAPS. Purple markers indicate ROIs selected by the full-data 82-ROI ElasticNet model but failing the 80% bootstrap stability threshold; gray markers indicate ROIs not selected by either criterion. Original model-development results, retained without refitting after the data audit.

Supplementary Figure S4

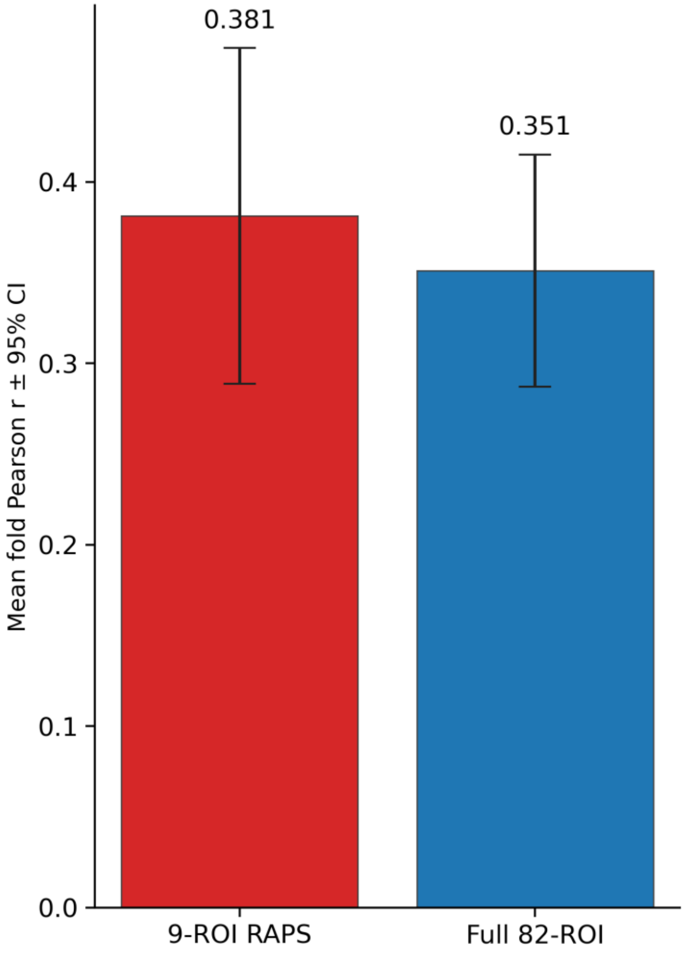

**Supplementary Figure S4 9-ROI RAPS vs Full 82-ROI Model Comparison.**

Comparison of post-selection cross-validated performance between the fixed 9-ROI RAPS model and the fully nested 82-ROI ElasticNet model, expressed as mean fold Pearson $r \pm$ 95% CI (empirical fold-level standard error from 5 outer folds). Mean fold correlations were numerically similar, but the validation designs differed because region selection was not nested for the nine-region model. Original model-development results, retained without refitting after the data audit.

Supplementary Figure S5

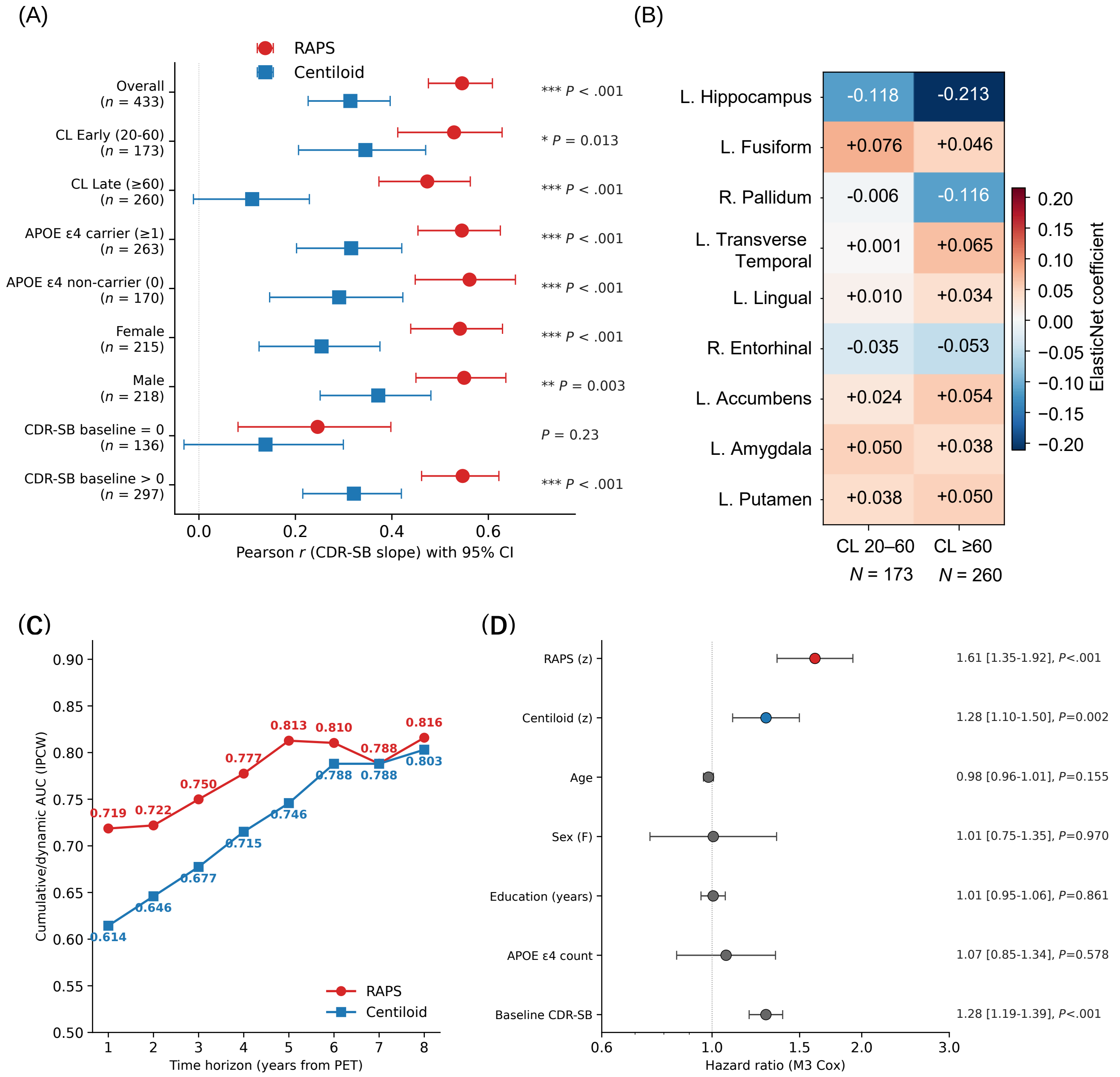

**Supplementary Figure S5 Subgroup Analysis and Clinical Utility.** (A) Forest plot of RAPS and Centiloid outcome correlations (Pearson *r* ± 95% CI; *P* = unadjusted Steiger test comparing RAPS vs CL; Benjamini-Hochberg-adjusted values in Supplementary Table S5) across subgroups: overall, CL Early (20–60)/Late (≥60), APOE ε4 carrier/non-carrier, sex, and cognitive status (CDR-SB = 0 vs CDR-SB > 0). (B) CL-stratified heatmap comparing ROI coefficients between early (CL 20–60) and late (CL ≥60) amyloid stages for the 9 RAPS ROIs. (C) Inverse-probability-of-censoring-weighted (IPCW) cumulative/dynamic time-dependent AUC over follow-up (1–8 years) for RAPS (red) and Centiloid (blue). At each time horizon, participants who had progressed by that time were classified as cases and participants observed to remain event-free beyond that horizon as controls; participants censored at or before the horizon were excluded from case-control pairs. RAPS shows higher discrimination than CL across early horizons (1–4 years), with convergence at later time points. (D) Cox proportional hazards forest plot showing hazard ratios with 95% CI for the fully adjusted M3 model including RAPS, CL, age, sex, education, APOE ε4 count, and baseline CDR-SB. RAPS and CL are scaled per SD; age and education are per year; APOE ε4 is per allele; baseline CDR-SB is per point; and sex compares female versus male. Panel B shows original model-development results, retained without refitting after the data audit.

Supplementary Figure S6

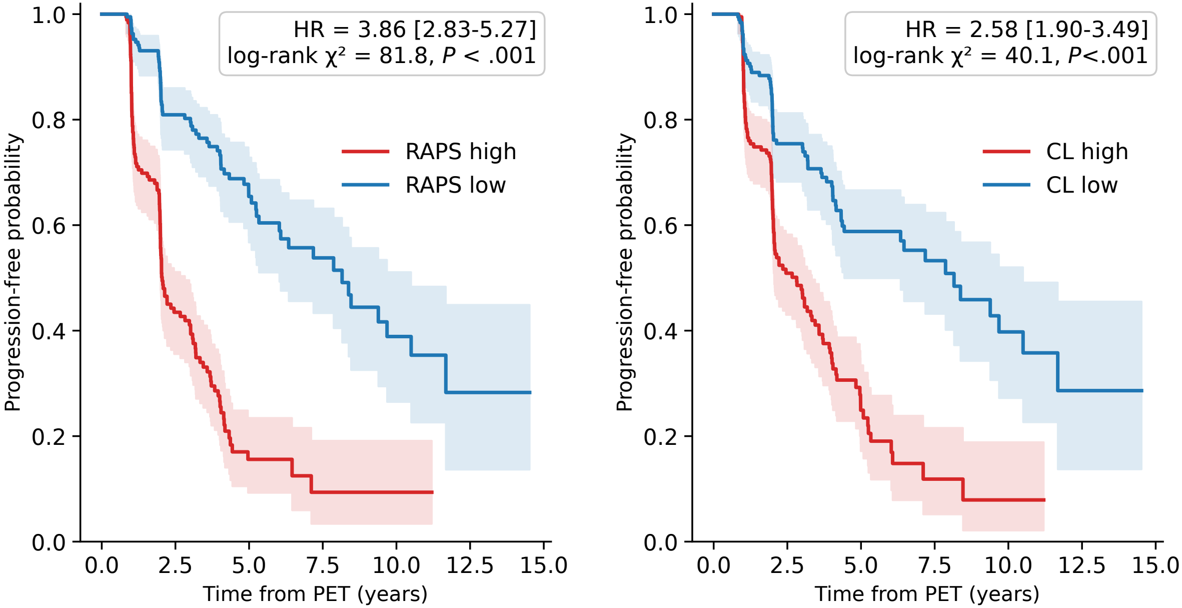

**Supplementary Figure S6 ADNI Cohort Survival Analysis.**

Kaplan-Meier survival curves for clinical progression (event = CDR-SB ≥ baseline + 1.0) in the ADNI discovery cohort, stratified by RAPS median split (*N* = 431; HR = 3.86, 95% CI 2.83–5.27; log-rank $\chi^2 = 81.8$, $P < .001$) and by baseline Centiloid median split (*N* = 431, as in the RAPS analysis; HR = 2.58, 95% CI 1.90–3.49; log-rank $\chi^2 = 40.1$, $P < .001$). Median time to clinical progression was 2.1 years in the RAPS-high group versus 8.2 years in the RAPS-low group.

Supplementary Figure S7

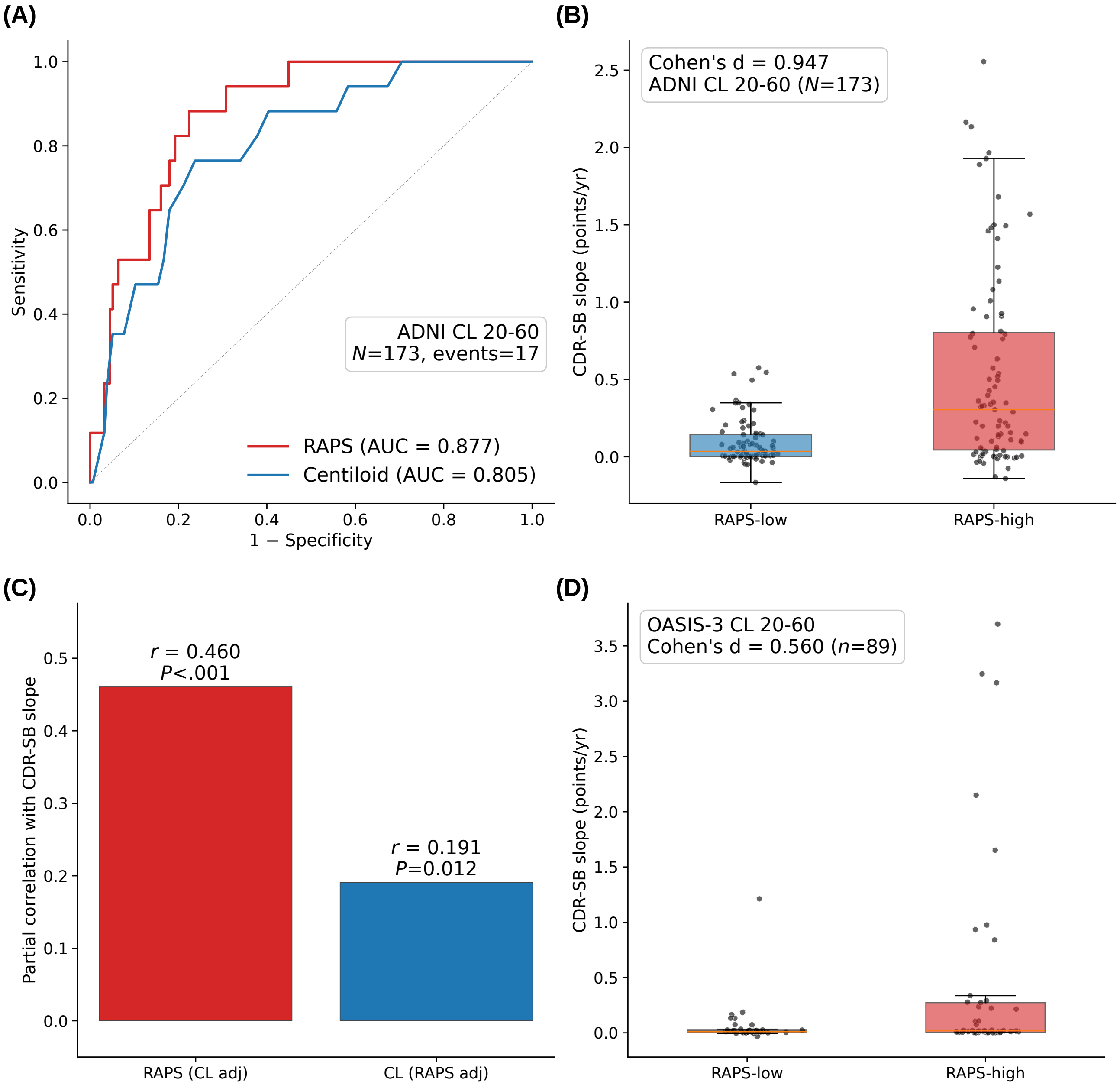

**Supplementary Figure S7 CL 20–60 Intermediate-Range Exploratory Analyses.** (A) ROC curves for discriminating rapid decliners (CDR-SB slope > 1.0/year) in the ADNI CL 20–60 subgroup ($N$ = 173), comparing RAPS and Centiloid. (B) RAPS-stratified CDR-SB slope distribution in ADNI CL 20–60 (RAPS-high vs RAPS-low median split, Cohen's $d$ = 0.947; boxplot fliers suppressed for clarity, individual subjects shown as overlaid points). (C) Partial correlation analyses for CDR-SB slope in ADNI CL 20–60: RAPS controlled for Centiloid versus Centiloid controlled for RAPS. RAPS retained a stronger independent association with CDR-SB slope than CL; CL showed a weaker residual association (CL controlled for RAPS: $r$ = 0.191, $P$ = .012). (D) OASIS-3 CL 20–60 subgroup boxplot of CDR-SB annual slope stratified by RAPS median split (Cohen's $d$ = 0.560, $n$ = 89), partially supporting the ADNI finding in an independent external cohort.

Supplementary Figure S8

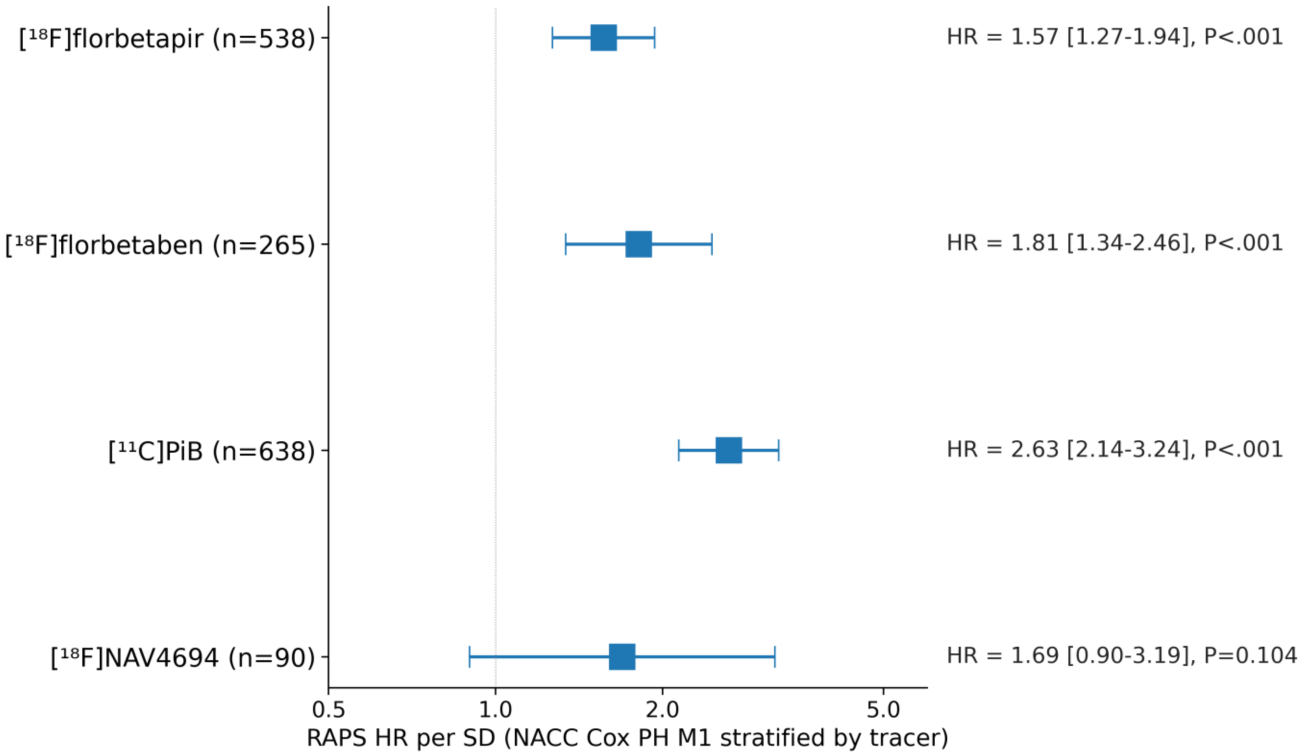

**Supplementary Figure S8 NACC SCAN Cross-Tracer Consistency Forest Plot.** Hazard ratios per SD of RAPS from multivariable Cox proportional-hazards models (M1: RAPS + age + sex), stratified by amyloid tracer in the NACC SCAN external validation cohort ($N$ = 1531). Hazard ratios are directionally consistent across [$^{18}$F]florbetapir (HR = 1.57, 95% CI 1.27–1.94, $P$ < .001; $n$ = 538, 79 events), [$^{18}$F]florbetaben (HR = 1.81, 95% CI 1.34–2.46, $P$ < .001; $n$ = 265, 27 events), and [$^{11}$C]PiB (HR = 2.63, 95% CI 2.14–3.24, $P$ < .001; $n$ = 638, 65 events). [$^{18}$F]NAV4694 (HR = 1.69, 95% CI 0.90–3.19, $P$ = .10; $n$ = 90, 7 events) is directionally consistent but exhibits a wide confidence interval reflecting limited statistical power from few events.

Supplementary Figure S9

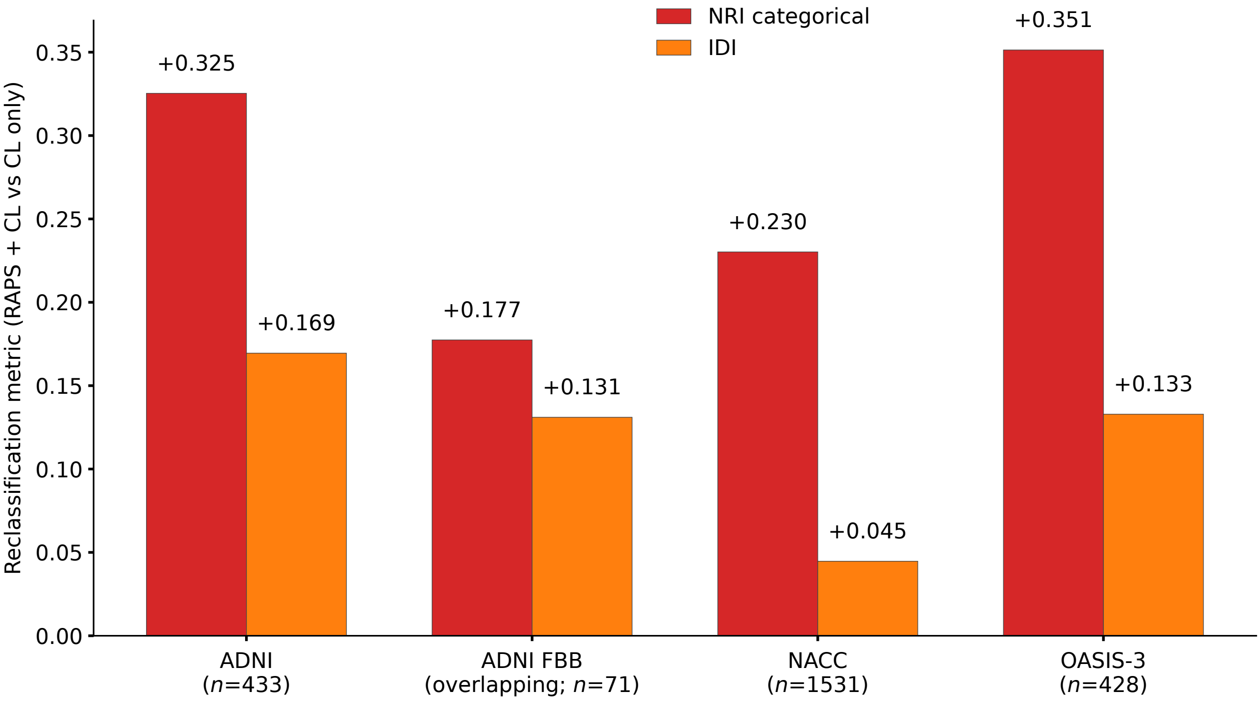

**Supplementary Figure S9 RAPS Reclassification Metrics.** Reclassification metrics (categorical net reclassification improvement [NRI] and integrated discrimination improvement [IDI]) quantifying the incremental discrimination of RAPS over Centiloid alone, computed across the ADNI discovery cohort, the overlapping ADNI FBB sensitivity subgroup, and the two external validation cohorts at the primary CDR-SB slope threshold (> 1.0/year). Values in event-limited external cohorts should be interpreted as exploratory. Positive values indicate favorable net reclassification or improved average discrimination relative to the Centiloid-only model; these estimates are exploratory and descriptive. IDI values are shown on the original probability scale.

Supplementary Figure S10

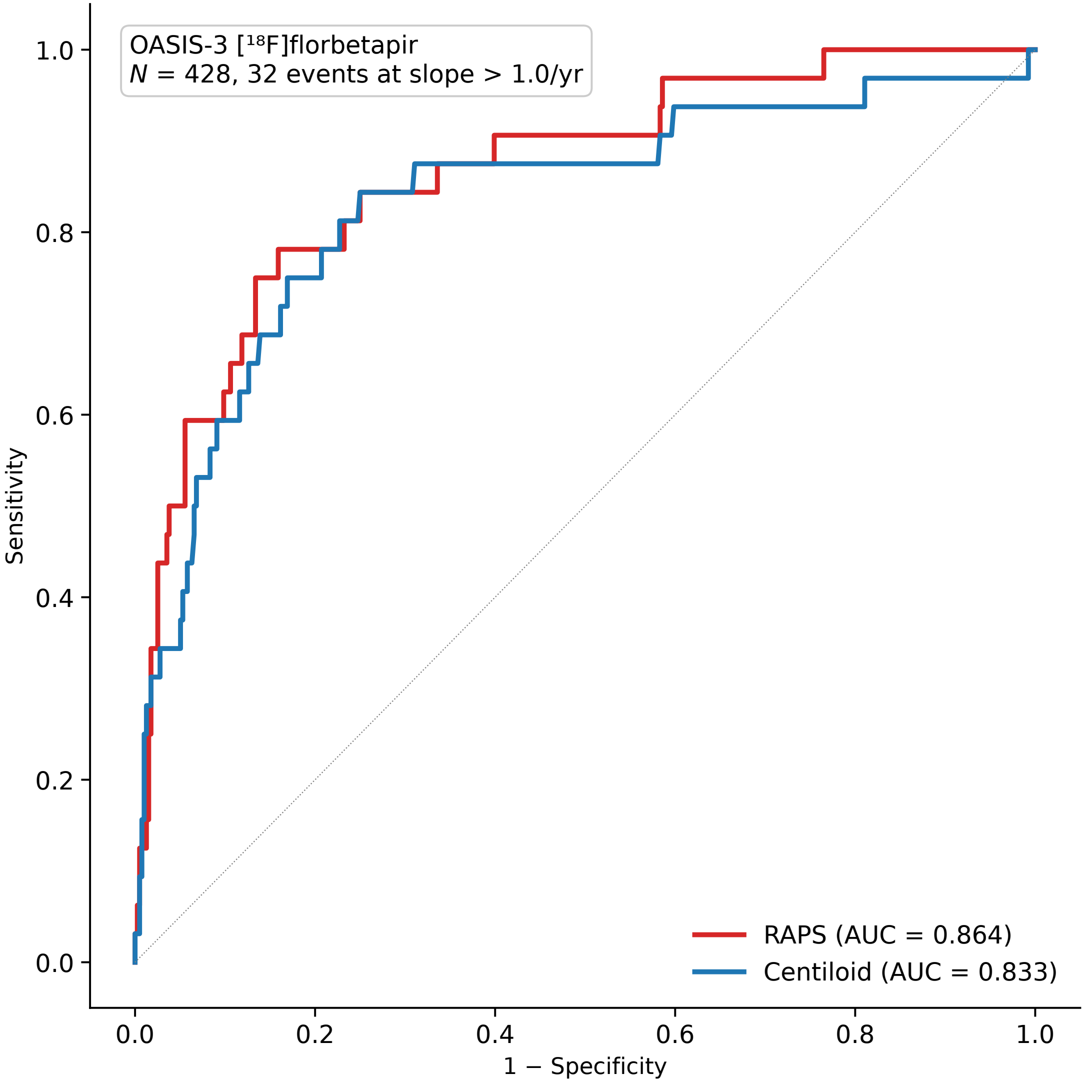

**Supplementary Figure S10 OASIS-3 ROC Curves.** Receiver operating characteristic curves for predicting rapid cognitive decline (CDR-SB slope > 1.0/year) in the OASIS-3 [$^{18}$F]florbetapir external validation cohort (*N* = 428 with available Centiloid), comparing RAPS and Centiloid. AUC values are shown in each curve legend.
